# Effects of Resistant Starch on Symptoms, Fecal Markers and Gut Microbiota in Parkinson’s Disease – The RESISTA-PD Trial

**DOI:** 10.1101/2021.02.07.21251098

**Authors:** Anouck Becker, Georges Pierre Schmartz, Laura Gröger, Nadja Grammes, Valentina Galata, Hannah Philippeit, Jacqueline Weiland, Nicole Ludwig, Eckart Meese, Sascha Tierling, Jörn Walter, Andreas Schwiertz, Jörg Spiegel, Gudrun Wagenpfeil, Klaus Faßbender, Andreas Keller, Marcus M. Unger

**Author notes:** **Corresponding author**: E-Mail (Unger MM), Department of Neurology, Saarland University, Homburg, 66421, Germany. Equal contribution (Anouck Becker, Georges Pierre Schmartz, Laura Gröger, Andreas Keller, Marcus M. Unger).

## Abstract

The composition of the gut microbiome is linked to multiple diseases, including Parkinson’s disease (PD). Bacteria producing short-chain fatty acids (SCFAs) and fecal SCFA concentrations are reduced in PD. SCFAs exert various beneficial functions in humans. In the interventional, monocentric, open-label clinical trial RESISTA-PD (NCT02784145) we aimed at altering fecal SCFAs by an 8-week prebiotic intervention with resistant starch (RS). We enrolled 87 subjects in three study-arms: 32 PD patients receiving RS (PD + RS), 30 control subjects receiving RS, and 25 PD patients receiving solely dietary instructions. We performed paired-end 100 base pair length metagenomic sequencing of fecal samples using the BGISEQ platform at an average of 9.9 GB. RS was well-tolerated. In PD + RS, fecal butyrate concentrations increased significantly and fecal calprotectin concentrations dropped significantly after 8 weeks of RS. Clinically, we observed a reduction in non-motor symptoms load in PD + RS. The reference-based analysis of metagenomes highlighted stable alpha-diversity and beta-diversity across the three groups, including bacteria producing SCFAs. Reference-free analysis suggested punctual, yet pronounced differences in the metagenomic signature in PD + RS. RESISTA-PD highlights that a prebiotic treatment with RS is safe and well-tolerated in PD. The stable alpha-diversity and beta-diversity alongside altered fecal butyrate and calprotectin concentrations calls for long-term studies, also investigating whether RS is able to modify the clinical course of PD.

## Introduction

Gut microbiota composition is known to be altered in Parkinson’s disease (PD) [1–3]. An increased abundance of *Enterobacteriaceae* has been consistently described in fecal samples of PD patients, whereas the abundance of *Prevotella, Faecalibacterium, Blautia*, and *Bifidobacterium* is reduced in PD [1,4–8]. This is of potential relevance, since bacteria with anti-inflammatory properties (e.g. synthesis of short-chain fatty acids, SCFAs) are less abundant, while potentially pro-inflammatory bacteria (e.g. endotoxin containing species) are more abundant in PD. Members of the family *Prevotellacae, Ruminococcacae*, and *Bacteroidacae* are capable of fermenting resistant starch (RS), a nutritional component which arrives in the large intestine without previous degradation by human enzymes [9]. Anaerobic fermentation of RS results in SCFAs, such as butyrate [10]. Butyrate exerts essential functions in the gut: it represents the main energy source for enterocytes, enhances gut motility, and exerts immunomodulatory effects [9,10]. Animal studies have shown that butyrate interacts with colonic regulatory T-cells, creating an anti-inflammatory environment [11]. Consequently, a lack of SCFA-producing bacteria and reduced colonic SCFA concentrations presumably lead to reduced gut motility as well as to a shift in the intestinal immune system toward a more pro-inflammatory environment [12]. Intestinal inflammation as well as altered gut motility (e.g. constipation) have frequently been described in PD. In addition, we have previously shown that PD patients have reduced fecal SCFA concentrations compared to matched controls [6].

With regard to techniques used to characterize the microbiome, 16S amplicon sequencing has been most frequently used in microbiome studies due to its broad availability, the moderate costs and the straightforward analysis. In recent years, whole genome sequencing (WGS) has become widespread available. Compared to 16S amplicon sequencing, WGS requires more complex computational and analytical procedures, but is superior in characterizing the metagenomic landscape with regard to resolution, accuracy, and functional profiling [13,14]. To characterize the metagenomic landscape, two different approaches can be used: Reference-free approaches characterize the metagenomic landscape based solely on sequencing data. Reference-based approaches rely on existing databases to compare the generated sequences against. In the present study, we computed the taxonomic profile with reference-based approaches. In addition, we also performed a comparative analysis with a hybrid approach named *BusyBee* [15], a software combining both (i.e. reference-free and reference-based) approaches.

A sensitive and valid marker of intestinal inflammation is fecal calprotectin. Calprotectin is a protein in human leukocytes. In case of inflammation, leukocytes migrate into the intestinal lumen and calprotectin can be measured in the feces as a stable marker that reflects even subclinical intestinal inflammation [16]. In accordance with the finding of prevailing pro-inflammatory bacteria in PD, elevated fecal calprotectin concentrations have been described in PD, too [17,18].

A prebiotic approach to increase SCFA concentrations is nutritional supplementation with RS. The efficacy and tolerability of a 12-week intervention with RS has already been shown in a controlled clinical trial for elderly subjects (≥ 70 years): RS was well-tolerated and, compared with placebo, elderly subjects on RS showed an altered intestinal microbiota, an increase in fecal butyrate concentrations, and a significant reduction in the use of laxatives [19].

Taken together, we set up the following hypothesis concerning a sequence of events: oral supplementation with RS enhances SCFA synthesis in the gut, probably accompanied by a shift in gut microbiota composition (due to a survival advantage for bacteria capable of fermenting RS). Consequently, the increased SCFA concentrations should lead to improved gut motility (improved constipation respectively) and a reduction in markers of intestinal inflammation.

## Results

### The RESISTA-PD study cohort

87 subjects participated in the RESISTA-PD trial. The study design and workflow illustrating subjects’ allocation to study-arms, clinical visits, sample collection, and analysis is summarized in **Figure 1**. The majority of subjects (n = 76) completed the study per protocol. Median age was 64.5 years in the PD group receiving RS (PD + RS), 66 years in the PD group receiving dietary instructions (PD + DI) group and 61.5 years in the control group receiving RS (Co + RS). There was no significant difference regarding sex ratio between the groups. The majority of subjects was on an omnivorous diet. Additional epidemiologic and clinical data are summarized in **Table 1**, detailed information regarding medication of the enrolled subjects is provided in Supplementary Table 1. No major side effects were reported during the 8-week intervention with RS.

**Table 1.** Epidemiological and clinical characteristics of the enrolled subjects

|  | **Parkinson patients**<br>**+ resistant starch** | **Control subjects**<br>**+ resistant starch** | **Parkinson patients**<br>**+ dietary instructions** |
| --- | --- | --- | --- |
| Number of subjects enrolled | n = 32 | n = 30 | n = 25 |
| Reasons to withdraw from study by subject's request after having started intervention | - Subject disliked taste of RS (n = 1)<br>- General discomfort and upset stomach after starting RS (n = 3)<br>- Concurrent acute disease after baseline visit (n = 1) | - Acute disease of family member (n = 1) | - Difficulties in handling fecal sampling kits at home (n = 1)<br>- Claiming "personal problems and family issues" (n = 1) |
| Number of subjects with 4-week follow-up | n = 28 | n = 29 | n = 23 |
| Number of subjects with 8-week follow-up | n = 26 | n = 27 | n = 23 |
| Age (median, [range]) | 64.5 [42 - 84] | 61.5 [40 - 76] | 66 [47 - 80] |
| Sex (male / female) | 18 / 14 | 12 / 18 | 13 / 12 |
| Dietary habits | Omnivorous diet: n = 29<br>Vegetarian diet: n = 2<br>Pescetarian diet: n = 1 | Omnivorous diet: n = 27<br>Vegetarian diet: n = 2<br>Pescetarian diet: n = 1 | Omnivorous diet: n = 25<br>Vegetarian diet: n = 0<br>Pescetarian diet: n = 0 |
| Smoker | 2 of 32 | 3 of 30 | 1 of 25 |
| Disease duration in months (median, [range]) | 111 [7 - 288] | not applicable | 111 [22 - 265] |
| History of appendectomy | 15 of 32 | 7 of 30 | 13 of 25 |
| UPDRS I, II, III total score in on state (median, [range]) | 35 [4 - 74] | not applicable | 30 [3 - 69] |
| MMSE (median, [range]) | 29 [23 - 30] | 30 [28-30] | 29 [25 - 30] |

*Note:* UPDRS, Unified Parkinson Disease Rating Scale; MMSE, Mini Mental State Examination.

**Figure 1.**
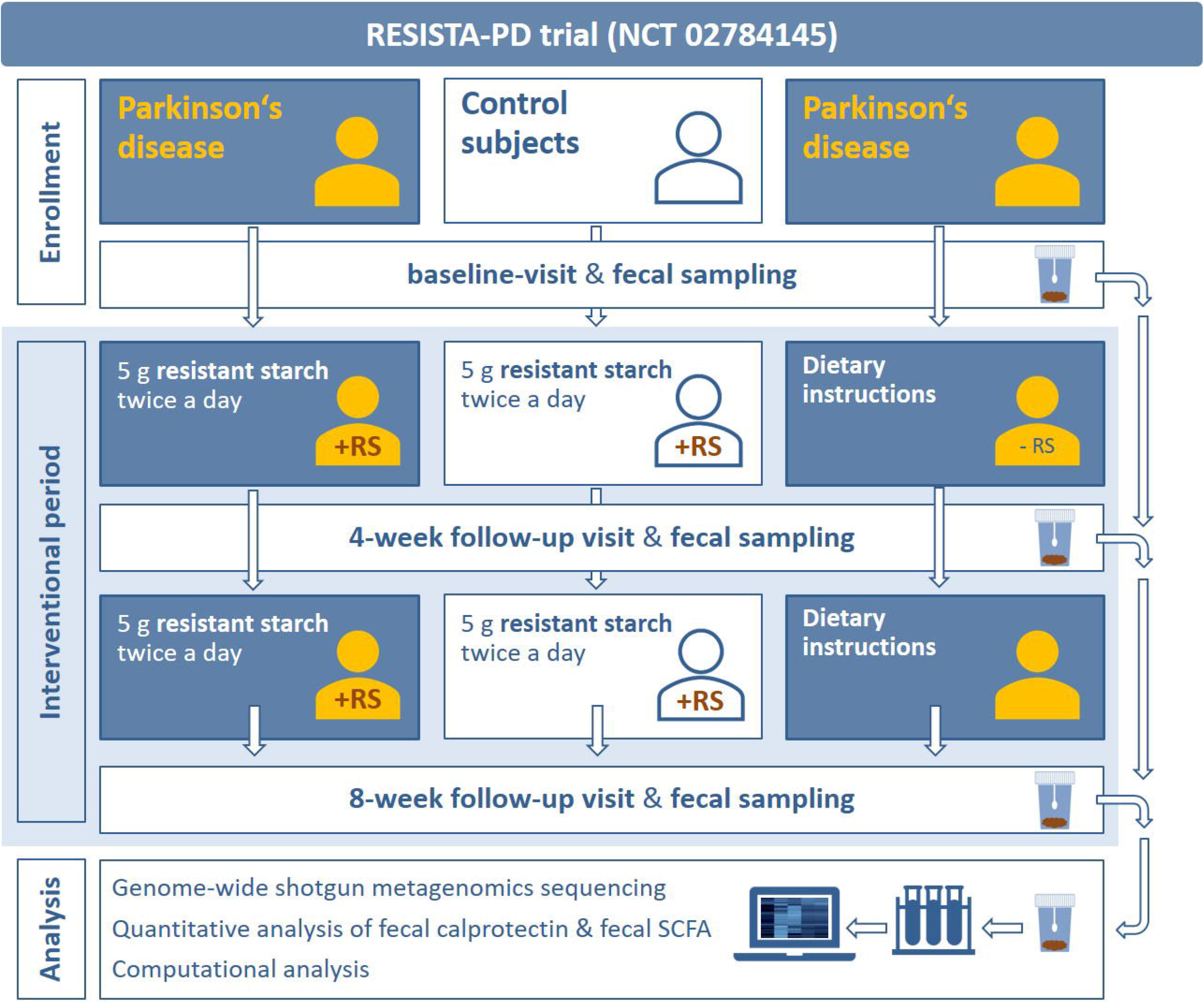
Study design. Subjects were assigned to three different study-arms. One group of Parkinson patients and a control group received 5 g resistant starch twice a day for a total period of 8 weeks. A second group of Parkinson patients solely received dietary instructions. Fecal samples and clinical scores were collected at baseline, after 4 weeks, and after 8 weeks for analysis. + RS in the pictograms visualizes subjects receiving resistant starch, -RS in the pictograms visualizes subjects not receiving resistant starch.

### Gut microbiota composition differs between PD patients and controls at baseline

At baseline, PD patients (n = 57) and controls (n = 30) showed no significant difference with regard to alpha-diversity with neither of the two applied analytical tools (*MetaPhlAn2* and *mOTUs2*) (Supplementary Figure 1A and 1B). With regard to beta-diversity, we observed a significant difference between PD patients and controls (p 0.001) with both analytical tools applied in this study (Supplementary Figure 1C and 1D). With regard to specific taxa, *Lachnospiraceae* species *incertae sedis* (mOTU_v25_12240, p 0.017) and *Faecalibacterium prausnitzii* (mOTU_v25_06110, p 0.019) showed significantly reduced abundances after correction for multiple testing in PD patients compared to controls (Supplementary Table 2). **Figure 2** illustrates descriptive differences at different taxonomic levels between PD patients and controls prior to the intervention. Descriptively, taxa of the phylum *Firmicutes* showed higher abundances in controls (except for the class *Bacilli*), while most taxa of the phyla *Proteobacteria*, especially *Enterobacteriaceae*, were more abundant in PD.

**Table 2.** Scores on clinical scales at baseline and post intervention

|  | **Parkinson patients**<br>**+ resistant starch** |  | **Control subjects**<br>**+ resistant starch** |  | **Parkinson patients**<br>**+ dietary instructions** |  |
| --- | --- | --- | --- | --- | --- | --- |
|  | Baseline | Post<br>intervention | Baseline | Post<br>intervention | Baseline | Post<br>intervention |
| CSS score<br>median [range] | 5 [0 - 14] | 3.5 [0 - 15] | 1 [0 - 11] | 0 [0 - 8] | 2 [0 - 10] | 2 [0 - 12] |
|  | p 0.257 |  | p 0.125 |  | p 0.674 |  |
| NMSQ score<br>median [range] | 10.5 [3 - 20] | 7.5 [2 - 18] | 3 [0 - 9] | 3 [0 - 10] | 10 [4 - 19] | 10 [5 - 19] |
|  | p 0.001 |  | p 0.774 |  | p 0.152 |  |
| BDI score<br>median [range] | 6.5 [2 - 25] | 3 [0 - 12] | 2 [0 - 14] | 2 [0 - 20] | 7 [1 - 18] | 6 [0 - 13] |
|  | p 0.001 |  | p 0.202 |  | p 0.106 |  |

*Note:* CSS, Constipation Scoring System; NMSQ, Non-Motor Symptoms Questionnaire; BDI, Beck Depression Inventory.

**Figure 2.**
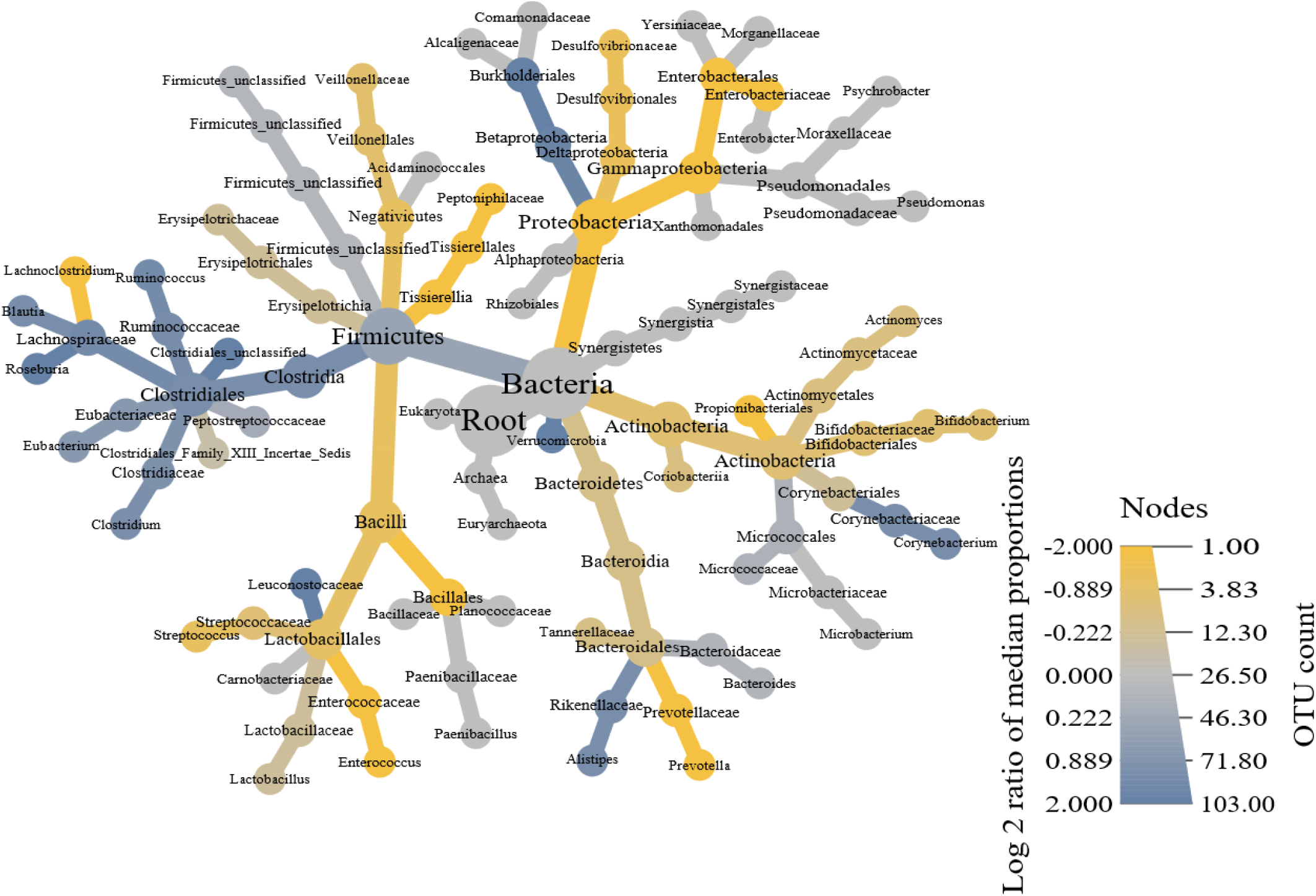
Taxonomic tree illustrating differences between PD patients and controls at baseline. Figure 2 shows a taxonomic tree illustrating the number of operational taxonomic unit (OUT) counts per taxon (visualized by the size of the radius) and the difference (visualized by color) between Parkinson’s disease (PD) patients and controls prior to intervention (baseline). Yellow shades indicate a higher abundance in PD patients, blue shades indicate a higher abundance in controls, grey shades indicate no group-specific differences. Low abundance taxa were pruned [47].

### Intervention with resistant starch alters symptoms load and fecal markers in PD

We next analyzed intervention-associated changes in subject-reported symptoms and in fecal markers. We observed a significant improvement with regard to non-motor symptoms (measured by the Non-Motor Symptoms Questionnaire score, p 0.001) and a significant improvement with regard to depressive symptoms (assessed by the Beck Depression Inventory, p 0.001) between baseline and 8 weeks in the PD + RS group (**Table 2**, Supplementary Figure 2a). No significant changes in these parameters were identified over the 8-week intervention period for PD + DI or Co + RS. There was no significant change in bowel habits (assessed with the Constipation Scoring System) between baseline and 8 weeks for any of the three investigated groups (**Table 2**, Supplementary Figure 2a). Calprotectin concentrations dropped significantly between baseline and 8 weeks (p 0.023, **Table 3**, Supplementary Figure 2b) in PD + RS. No significant changes in fecal calprotectin concentrations were observed between baseline and 8 weeks in the two other groups (i.e. Co + RS and PD + DI). Concerning fecal SCFAs, the concentration of the SCFA butyrate increased significantly in PD + RS between baseline and 8 weeks (for absolute fecal butyrate concentrations (p 0.029) as well as for relative fecal butyrate concentrations (p 0.026, **Table 4**, SupplementaryFigure 2c). There were no significant changes for SCFAs other than butyrate between baseline and 8 weeks in PD + RS and no significant changes regarding SCFA concentrations in the two other groups (i.e. Co + RS and PD + DI).

**Table 3.** Fecal calprotectin concentrations at baseline and post intervention

|  | **Parkinson patients**<br>**+ resistant starch** |  | **Control subjects**<br>**+ resistant starch** |  | **Parkinson patients**<br>**+ dietary instructions** |  |
| --- | --- | --- | --- | --- | --- | --- |
|  | Baseline | Post<br>intervention | Baseline | Post<br>intervention | Baseline | Post<br>intervention |
| Calprotectin<sup>1</sup><br>median [range] | 56.8 [19 - 327] | 20.5 [19 - 407] | 19 [19 - 69] | 19 [19 - 155] | 46 [19 - 219] | 31 [19 - 217] |
|  | p 0.023 |  | p 1.000 |  | p 0.481 |  |

<sup>1</sup>Calprotectin values are µg per g feces.

**Table 4.**
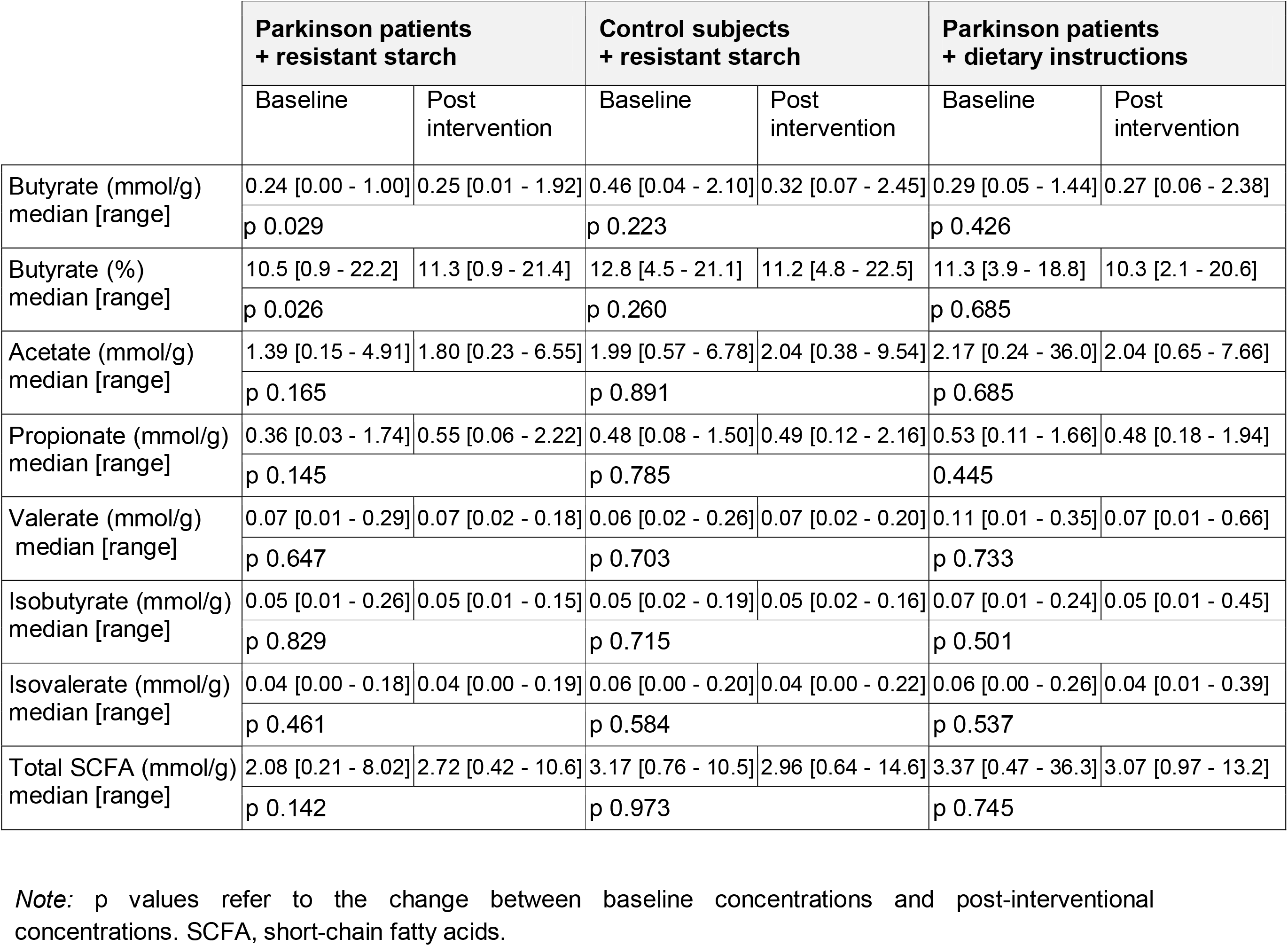
Short-chain fatty acid concentrations at baseline and post intervention

### Reference-based analysis shows a stable gut microbiome after RS intervention

In order to investigate whether the observed changes in clinical symptoms and fecal markers are associated with an intervention-associated shift in the gut microbiome, we performed metagenomic sequencing. Quality control by FastQC indicated good data quality of metagenomic sequencing. During preprocessing, less than one percent of reads was removed for each sample. Additionally to the standard quality control, we analyzed pairwise *Mash distances* [20] between all samples. Hereby, the *Mash distance* gauges similarity between sequencing libraries using only sequence features directly derived from raw reads. Visualizing *Mash distances* shows that samples derived from the same individual frequently produce the lowest *Mash distance*, indicating correct labeling of samples and a lack of contamination (**Figure 3**). No intervention-associated changes with regard to either alpha-diversity or beta-diversity were detected for any of the three investigated groups (PD + RS, PD + DI, Co + RS). No significant intervention-associated changes were also detected concerning differences in distinct taxa (Supplementary Table 2). Non-metric multidimensional scaling (NMDS) visualizing microbiome shifts did not reveal uniform shifts associated with the intervention (Supplementary Figure 3). **Reference-free analysis points at punctual differences in the metagenomic signature** Reference-free analysis revealed intervention-associated changes in taxonomic signatures in PD + RS (**Figure 4**). The majority (>54%) of contigs forming one of the three clusters in the reference-free analysis derived from the genus *Rhodococcus* (Supplementary Figure 4). Density changes worth interpreting as clusters identified in the other cohorts (Co + RS and PD + DI) did not contain significant amounts of *Rhodococcus* sequences.

**Figure 3.**
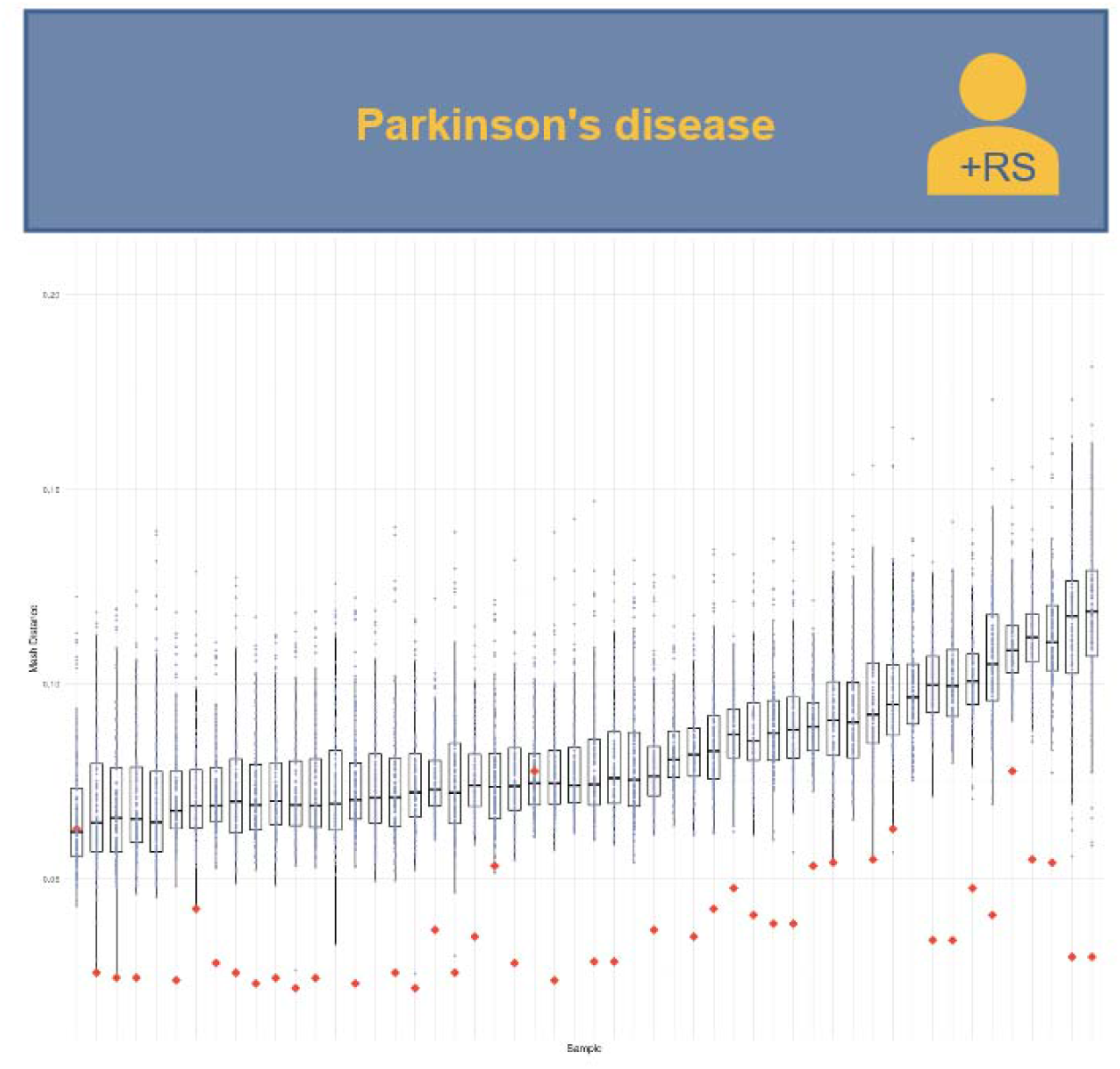
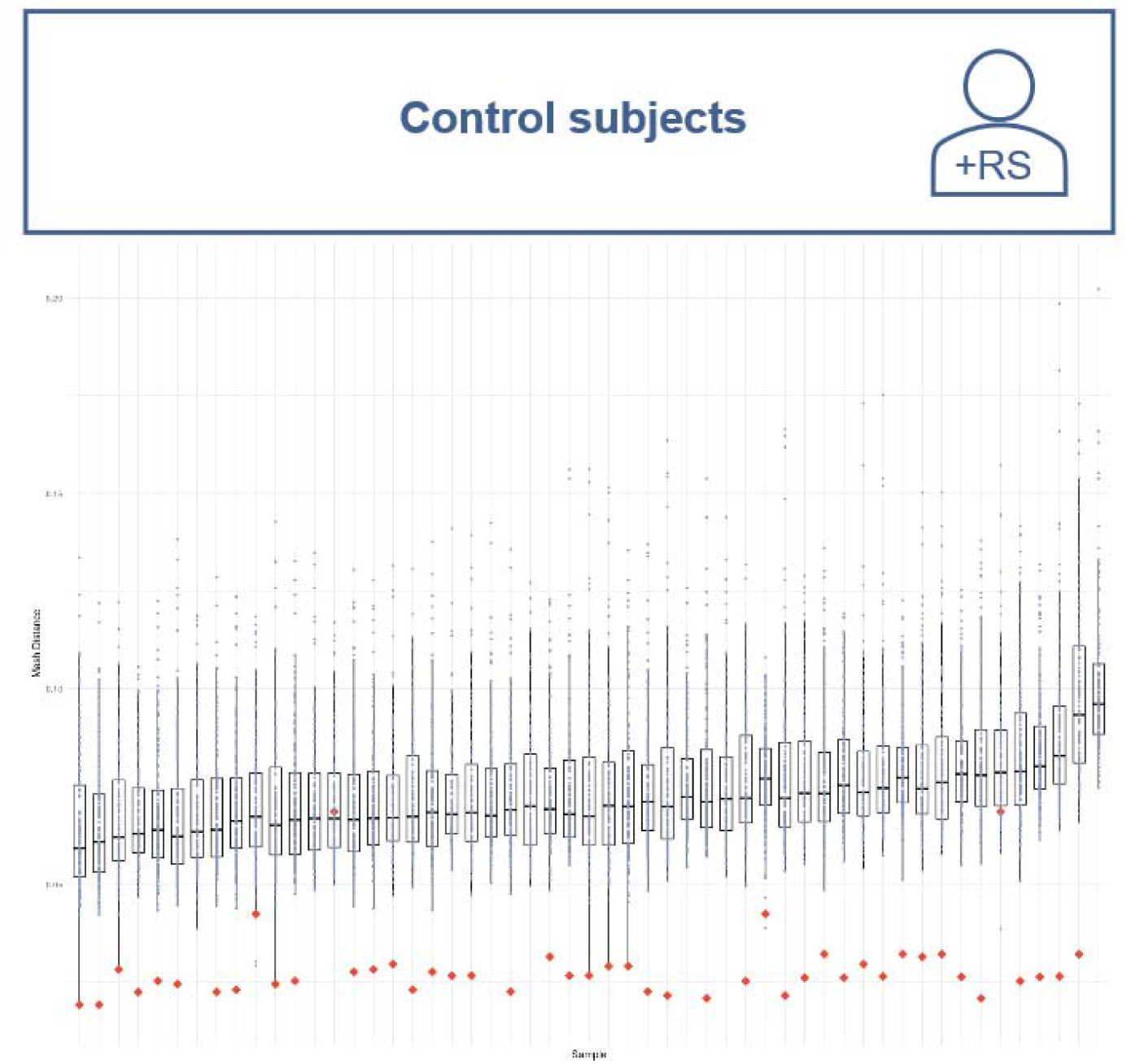
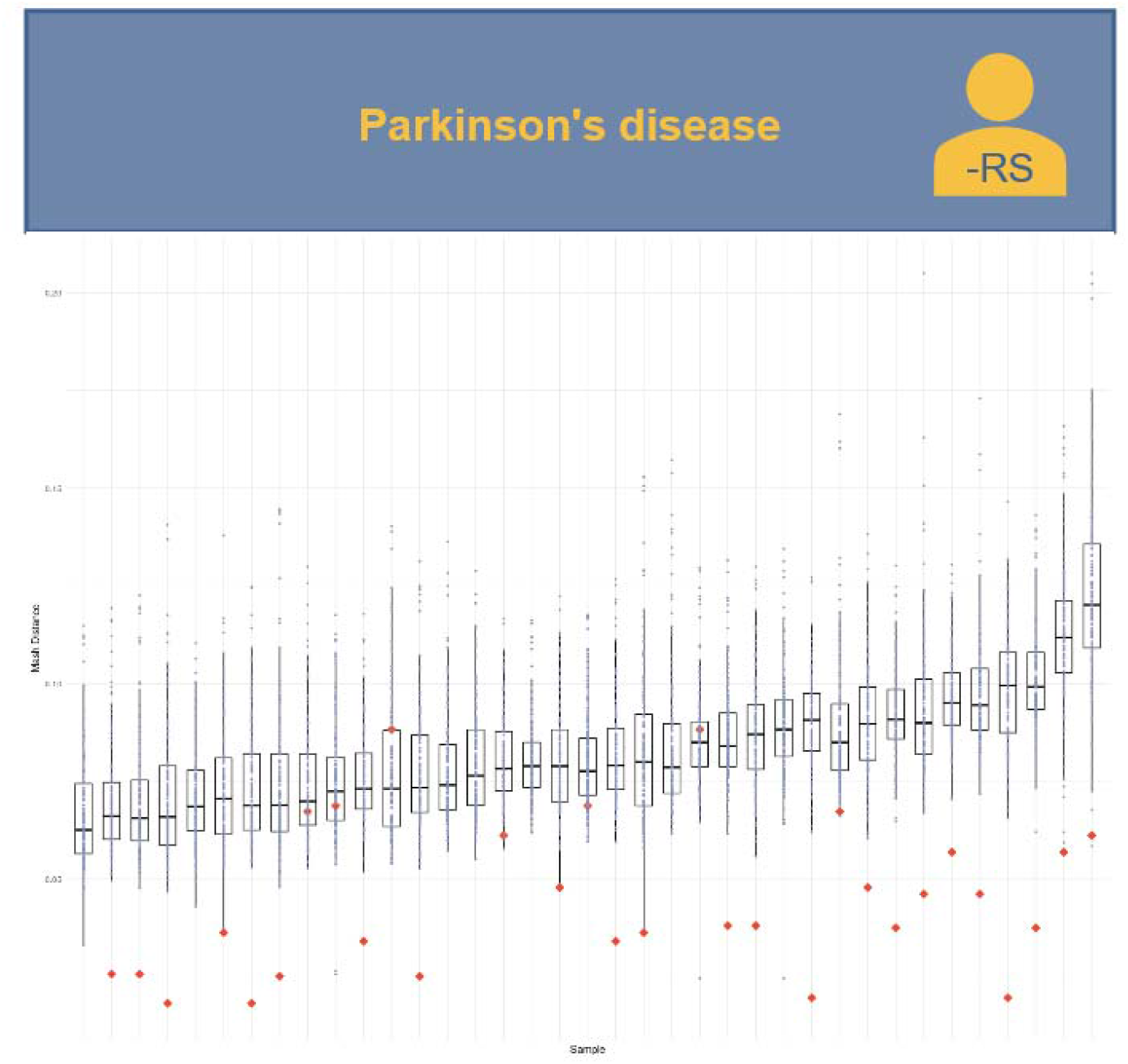
Intra-individual and inter-individual similarity of samples. Similarity of samples visualized as *Mash distance* plot. The lower the *Mash distance*, the higher the similarity of samples. Red diamonds represent paired samples (baseline and 8 weeks) of one subject. Dots represent samples of other subjects (unpaired). **A** Parkinson’s disease patients receiving resistant starch (+ RS), **B** control subjects receiving RS (+ RS) and **C** Parkinson’s disease patients receiving solely dietary instruction, but no resistant starch (-RS).

**Figure 4.**
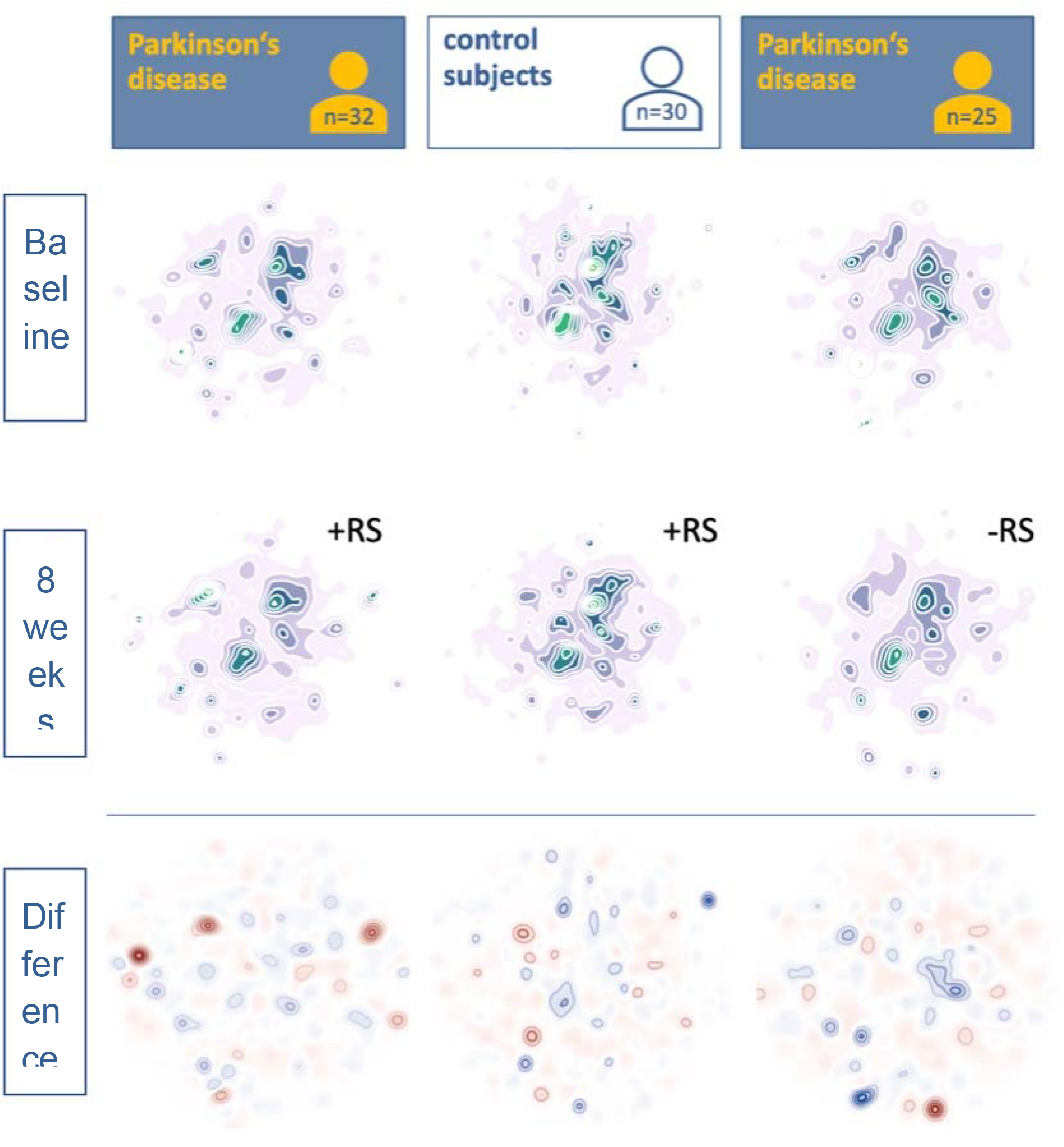
Reference-free analysis points at punctual differences. Figure 4 shows the density distribution of the 5-mers, after dimensionality reduction with uniform manifold approximation and projection. The first row contains the baseline. The second row shows the 8-week follow-up. In the bottom row the difference between the two previous rows is visualized; blue indicates a stronger signal at baseline, red indicates a stronger signal at 8 weeks follow-up. + RS indicates the two groups receiving resistant starch, -RS indicates the group not receiving resistant starch.

### Distinct microbial signatures are associated with fecal butyrate concentrations

In metagenomic samples, the change in abundance of one taxon is likely to entail changes in the abundance of other taxa. We investigated our data for data compositionality using the *selbal* algorithm. *Selbal* searches for two groups of taxa whose relation (or balance) is associated with a certain response variable. The relationship is modeled as linear or logistic regression model of the taxa on the response variable. *Selbal* builds multiple models containing different taxa combinations and evaluates their performance using cross-validation. In our dataset, response variables were measurements of acetate, propionate, butyrate, valerate, calprotectin, as well as CSS and BDI scores. Using the *selbal* algorithm, results for *MetaPhlAn2* (**Figure 5A and 5B**) and for *mOTUs2* data (**Figure 5C and 5D**) and butyrate concentrations as response variable were highly consistent. For absolute butyrate concentrations, *selbal* detected that higher abundances of *Fusicatenibacter saccharivorans* to *Ruthenibacterium lactatiformans* were associated with higher absolute butyrate concentrations (**Figure 5A**) with an association slightly below moderate (*MetaPhlAn2* data, R = 0.35). The association of *Ruthenibacterium lactatiformans* with butyrate concentrations was verified by *mOTUs2* data (**Figure 5C**). Here, *selbal* detected that a higher abundance of *Lachnospiraceae* and *Streptococcus parasanguinis* to *Ruthenibacterium lactatiformans* was associated with higher absolute butyrate concentrations. The association was moderate (R = 0.44). For relative butyrate concentrations, *selbal* detected that a higher abundance of *Dorea longicatena* (*MetaPhlAn2* data) and *Blautia wexlerae* (*MetaPhlAn2* and *mOTUs2* data) with respect to *Ruthenibacterium lactatiformans* (*MetaPhlAn2* and *mOTUs2* data) was associated with higher butyrate concentrations (**Figure 5B and 5D**). The association was moderate (R = 0.48 for *MetaPhlAn2*, R = 0.5 for *mOTUs2*). The model itself was stable, with *Ruthenibacterium lactatiformans* and *Dorea longicatena* being included in over 95% of all models (*MetaPhlAn2*) and *Blautia wexlerae* and *Ruthenibacterium lactatiformans* being included in over 96% of all models (*mOTUs2*). Other response variables did not show a consistency between *mOTUs2* and *MetaPhlAn2* data.

**Figure 5.**
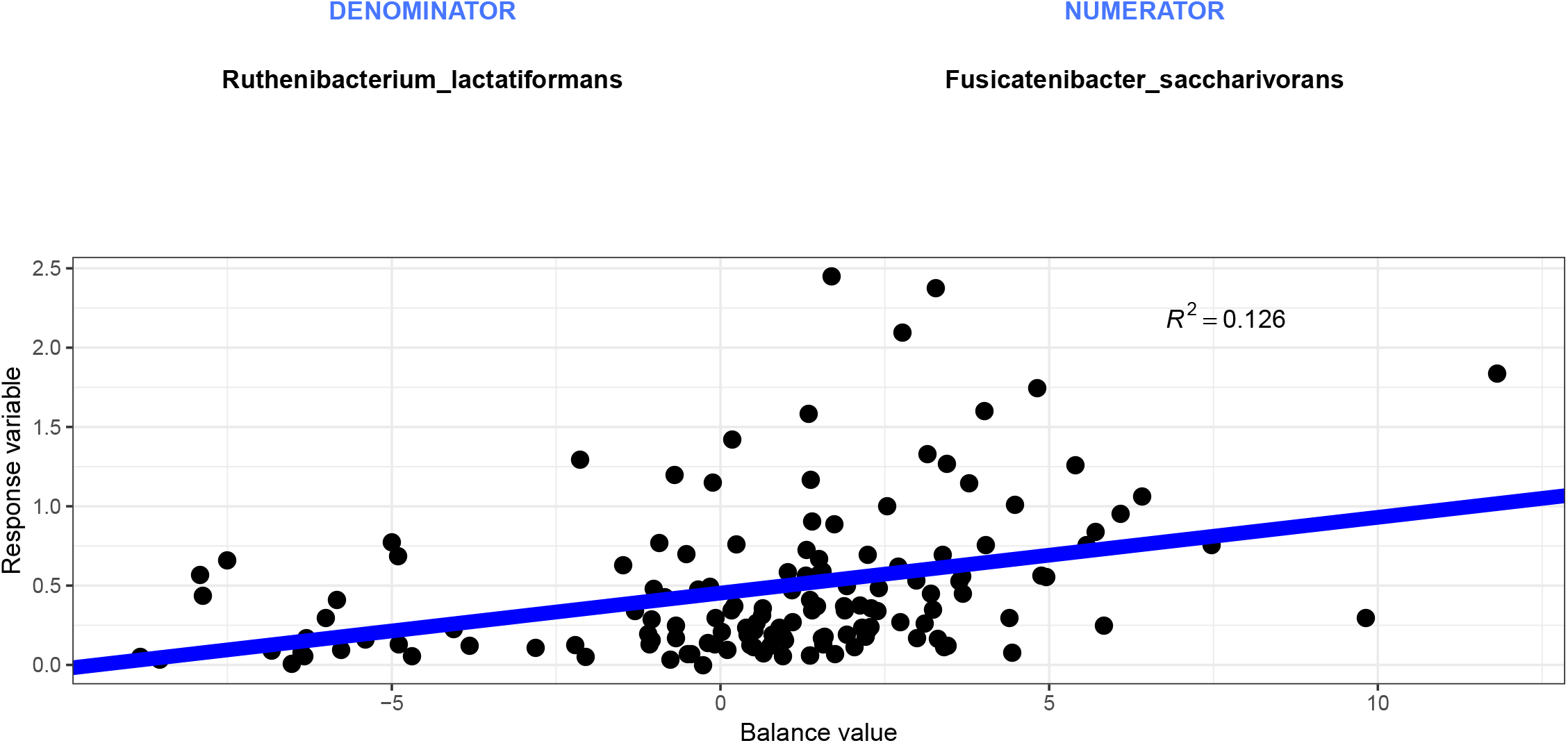
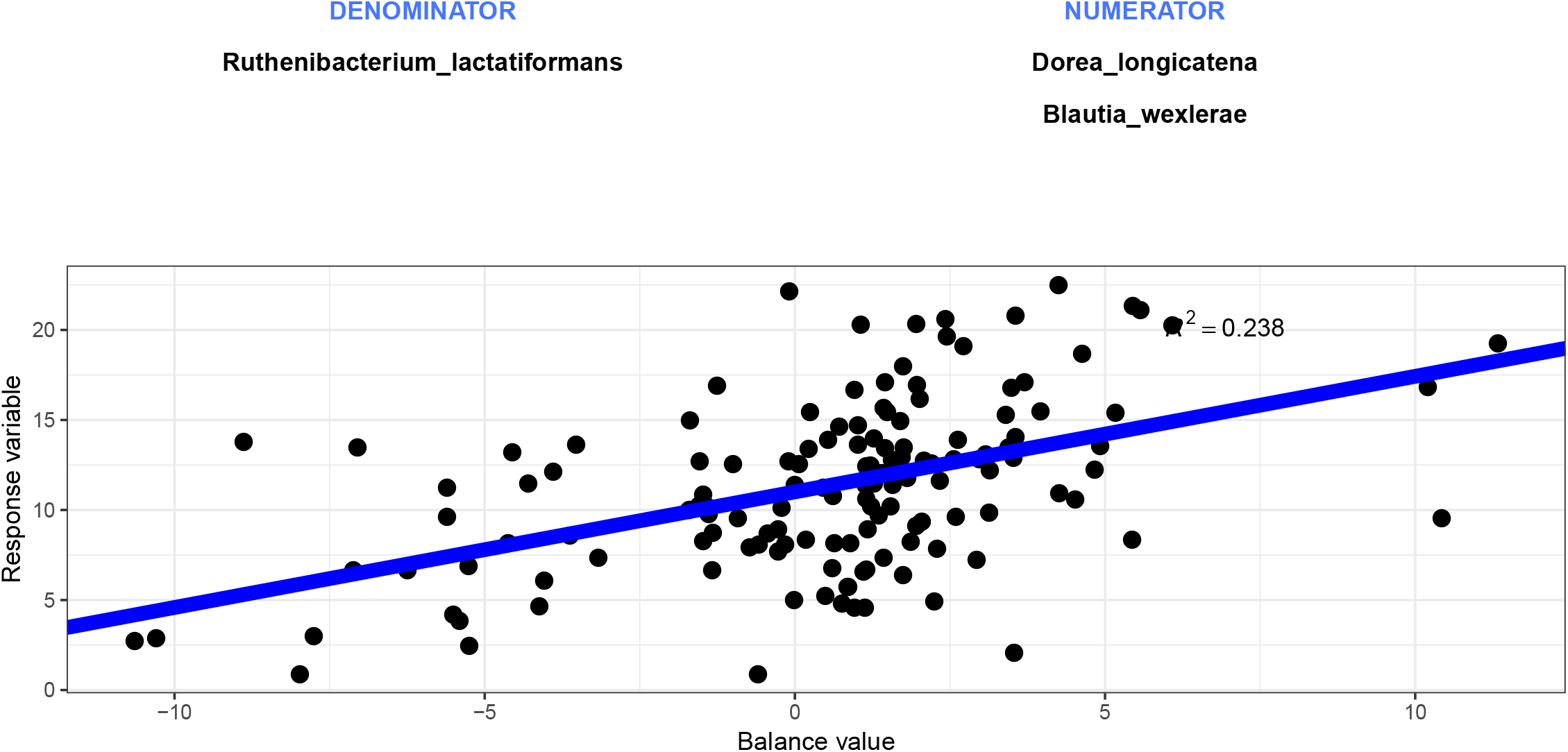
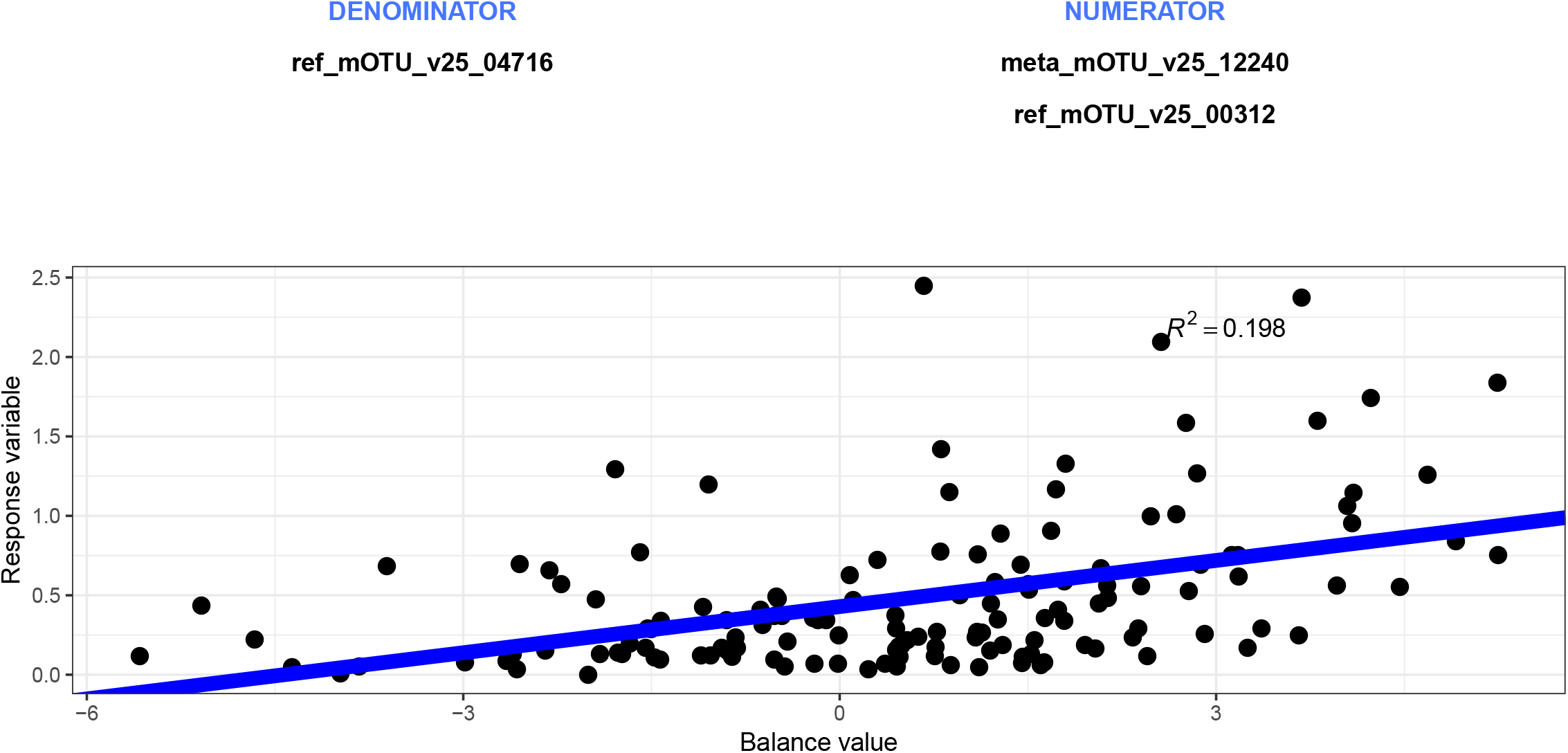
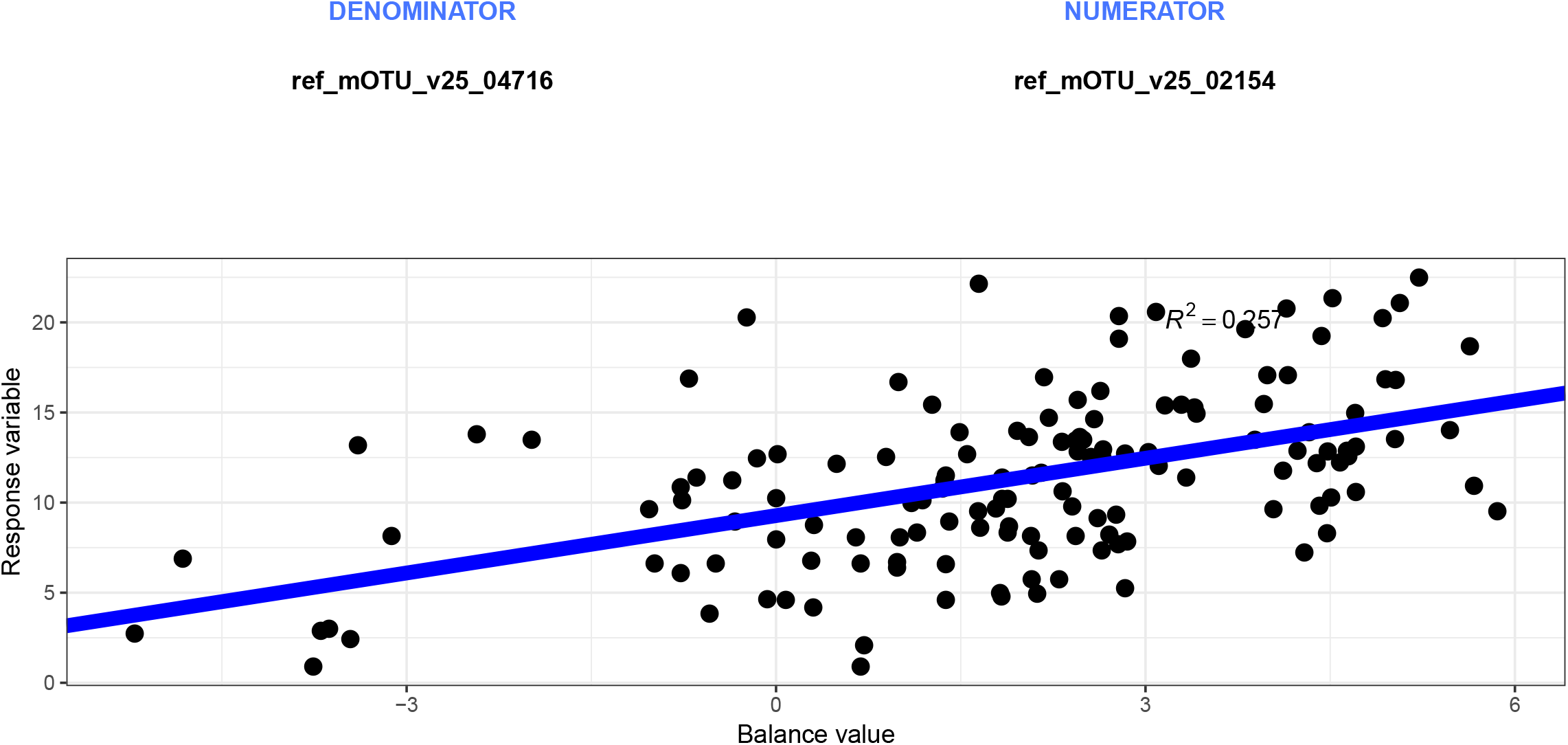
Distinct microbial signatures are associated with fecal butyrate concentrations. Balance scores for *MetaPhlAn2* (5A, 5B) and *mOTUS2* (5C, 5D) data with absolute butyrate concentrations (5A, 5C) and relative butyrate concentrations (5B, 5D) as response variables.

### Functional profiling reveals no intervention-associated difference

In order to identify differences in the available metabolic pathways, we applied the *HUMAnN2* tool to our data. The estimated pathway abundances were used for an exploratory data analysis of the samples using principal component analysis (PCA) and a differential analysis using *ALDEx2*. The PCA projection indicated a different tendency between the PD and the control groups, but no differences associated with the intervention (baseline vs. 8 weeks) (Supplementary Figure 5). The analysis with *ALDEx2* did not result in any pathway that showed a significant difference between groups nor a difference between baseline and 8 weeks (Supplementary Table 3).

## Discussion

Gut microbiota composition is altered in PD [3–6] and might be a contributing factor for gastrointestinal non-motor symptoms (e.g. constipation) in PD. Having recognized the relevance of the intestinal microbiome in PD, probiotics have been investigated in PD and other neurodegenerative diseases previously [21,22] and prompted us to perform the RESISTA-PD trial.

In accordance with other studies in the field [2,4,5,7,23,24], we observed a difference between PD patients and controls at baseline regarding beta-diversity. With regard to specific taxa, we detected significantly different abundances for two taxa after correction for multiple testing: abundances for *Lachnospiraceae* species *incertae sedis* and *Faecalibacterium prausnitzii* were significantly reduced in the fecal samples of PD patients. *Lachnospiraceae* as well as *Faecalibacterium prausnitzii* have already been reported to be reduced in PD and have also been confirmed as altered taxa in PD in a recent meta-analysis [25]. Indeed, the lower abundance of *Faecalibacterium prausnitzii* might be one explanation for the lower fecal butyrate concentrations in PD. On a descriptive level, we also reproduced some other previously reported alterations of gut microbiota in PD, e.g. a lower abundance of *Firmicutes* and a higher abundance of *Proteobacteria*, especially *Enterobacteriaceae*.

For the reference-free analysis of intervention-associated changes, a metagenomic signature indicating a possible involvement of *Rhodococcus* was found in PD + RS, despite an insignificant change in abundance during the read-based analysis. The blue cluster (top right) in the CO + RS group contained only sequences derived from one single sample. Sequences were assigned to *Rhodotorula toruloides*. The two clusters with the highest density in the PD + DI group contained no sequences of the genus *Rhodococcus* (blue cluster) and less than 0.001 % of sequences (red cluster) of the genus *Rhodococcus*.

The reference-free workflow we selected discards all quantity information after assembly. In an optimal scenario, sequences derived from identical genetic information will be collapsed into the same contig in each sample. In *BusyBee* one such contig will appear as one individual point with close to no impact on the overall density distribution. The strong signal from the high-density cluster in PD + RS suggests the existence of multiple contigs that were dissimilar enough not to be collapsed during assembly, yet qualitatively good enough to be assigned to *Rhodococcus*. Accordingly, the change in density of the investigated cluster indicates a more complex behavior than a quantitative balance shift. Instead, an increase in genomic diversity may be postulated from this observation. The relevance of this particular finding remains unclear and requires further investigations, especially since the genus *Rhodococcus* is not a typical representative of the human gut microbiota. Given the fact that bacteria of the genus *Rhodococcus* are not typically part of the human gut microbiota and also are obligate aerobes, the identification of this genus in human fecal samples points at a potential contamination. Indeed, *Rhodococcus* has been identified in human biosamples due to DNA contamination of reagents [26]. Yet, DNA contamination is mainly a problem when analyzing low microbial biomass samples (like blood or saliva). In our study that analyzed high microbial biomass samples (feces), contamination is further unlikely as contamination would have occurred solely in PD + RS samples after the intervention and not in the other two groups. One might also hypothetically consider contamination of a single batch of reagents or tubes used in our study. However, samples were analyzed in a random way and not sorted by group prior to analysis. Hence, contamination can hardly explain this finding. As the taxonomic assignment of contigs relies on libraries, misassignment due to similar sequences of another taxon (not represented in libraries) with sequences of *Rhodococcus* should also be considered.

Our finding that a prebiotic intervention with RS significantly alters fecal butyrate concentrations and significantly reduces fecal calprotectin concentrations is also in line with a controlled clinical trial that investigated RS in mid-age and elderly subjects and reported an increase in fecal butyrate concentrations in subjects aged 70 years or older [19]. While the study by Alfa and colleagues [19] even indicated a therapeutic effect (reduction in the use of laxatives), we did not observe a significant improvement of bowel habits. This divergent observation between our study and the study by Alfa and colleagues might be due to different types of RS (we used RS type 3, Alfa and colleagues used RS type 2), the dose of RS (Alfa and colleagues administered approximately double the dosage compared to our study), and the duration of the interventional period (8 weeks in our study versus 12 weeks in the study by Alfa and colleagues).

Fecal butyrate concentrations and calprotectin concentrations were not altered when PD patients solely underwent nutritional counseling including dietary instruction concerning a fiber-rich diet (PD + DI). Considering the fact that the PD + DI group underwent the same visit schedule as the PD + RS group, the effect observed with regard to clinical measures in PD + RS, is unlikely to be completely due to unspecific effects such as attention paid to subjects during clinical visits or answering questionnaires according to social desirability.

The effect on symptoms related to depression in the PD + RS group might be explained by the observed increase in butyrate concentrations. An association between gut microbiota and depressive symptoms has been described previously [27, 28]. Administration of SCFA, including butyrate, has shown to reduce depressive symptoms in mice [29]. Moreover, fecal SCFA have been shown to be reduced in a cohort of female patients with depression [30]. Increasing evidence suggests a connection between depressive symptoms and fecal SCFA concentrations [31]. One explanation for the lack of a change in clinical measures in the PD + DI group might be that adherence to dietary instructions is likely to be lower compared to the more convenient approach of consuming a dietary supplement (dissolved in one glass of water) twice a day. In addition, changes in dietary habits are much more heterogeneous compared to a standardized nutritional supplementation.

The fact that Co + RS did not show a reduction in fecal calprotectin concentrations is likely to be explained by already normal calprotectin concentrations in control subjects at baseline. The unchanged SCFA concentrations in Co + RS might be either explained by a ceiling effect or by a lower adherence (as controls did not expect to benefit from the intervention).

Even though the effects on fecal calprotectin and fecal butyrate were significant in PD + RS and also SCFAs other than butyrate showed a trend towards an increase in concentrations in the PD + RS group, our data lack a clear-cut correlate concerning specific gut microbiota. Assuming that gut microbiota composition remained stable despite the prebiotic intervention, an altered transcription might have led to the observed effects on fecal markers. The lack of a clear-cut response to the intervention with regard to gut microbiota or symptoms of constipation might also be due to various individual factors. Our study design controlled for confounding factors like age, sex, overall type of diet, comorbidities, and medication. Nevertheless, the investigated cohorts were heterogeneous (even within groups) with regard to other, more complex factors that might determine the individual response (e.g. composition of the gut microbiome prior to the intervention, adherence to the recommended RS intake, more specific dietary habits). This said, the limited sample size in this proof of concept study together with the inter-individual variability concerning potential confounding factors is one explanation for the heterogeneous response to the intervention. Hence, larger cohorts (as well as transcriptomics and proteomics) might have been necessary to detect more subtle intervention-associated alterations in the gut microbiome (and possible changes at the transcriptomic level).

In order to identify microbial signatures associated with SCFA concentrations, we performed an *in silico* analysis (using the *selbal* algorithm): Balance analysis of taxa and butyrate concentrations resulted in concordant results for both analytical tools (*mOTUs2* and *MetaPhlAn2*). Moreover, we confirmed the robustness of the identified balance scores by their frequency in a cross-validation model. The microbial taxa *Blautia wexlerae, D. longicatena* and *Ruthenibacterium lactatiformans* are involved in butyrate-related pathways [32]. However, all of these bacteria are not capable of directly producing butyrate from RS, but they produce lactate and succinate by fermentation which consecutively serve as substrates for other bacteria which produce butyrate [10]. Despite the fact that our *in silico* approach did not detect classical SCFA producers (like *Faecalibacterium* or *Roseburia*) as determinants for fecal butyrate concentrations, the taxa identified by *selbal* are indirectly involved in butyrate production (via complex interactions with other taxa) [10].

In contrast to our initial hypothesis, symptoms related to constipation (a frequent non-motor symptom in PD) were not significantly altered during the 8-week-intervention. As there was at least a descriptive decline in CSS scores after RS supplementation, we suggest longer interventional periods and increased doses of RS to test such a symptomatic effect on bowel habits. Given that RS was well-tolerated in the RESISTA-PD trial, this seems to be a feasible and rational approach.

Besides the limited sample size and the relatively short interventional period, one main limitation of the RESISTA-PD trial is its open-label study design. We aimed at counteracting this shortcoming by including an additional PD control-arm (PD + DI) to control for unspecific effects (as discussed above). Adherence to the intervention was checked by patient diaries but not by more objective measures. Probably, internal motivation to adhere to the study protocol might have been higher in PD + RS compared to Co + RS (as discussed above). The primary aim of the RESISTA-PD trial was to test the feasibility, tolerability and efficacy of this prebiotic approach. Hence, our study protocol did not include an additional measurement of the investigated markers several weeks after withdrawal of RS. We suggest to include such an assessment in future studies.

At this time, we are not able to answer the question whether the observed anti-inflammatory effects indicated by the decline in fecal calprotectin concentrations is mediated by the increase in butyrate concentrations. Even though other studies endorse such an assumption [11], further studies are needed to clarify the exact mechanisms of this prebiotic intervention and the increase in SCFAs in details.

A general limitation of interventions aiming at altering the gut microbiome is the question of endurance. This is why long-term studies and assessment of subjects after withdrawal of the intervention are mandatory to draw final conclusions.

## Conclusions

RS, as a dietary supplement to increase fiber intake, is safe and well-tolerated in PD. RS supplementation partially restores fecal SCFA concentrations in PD + RS without clear-cut changes in the gut microbiome that were attributable to the intervention. Alterations at the transcriptome level (that were not captured by our approach) might explain the intervention associated significant increase in fecal markers in the PD + RS group.

In view of the good tolerability of RS, we suggest long-term studies with RS. These studies should also aim at clarifying the underlying mechanisms for the supposed anti-inflammatory effects. Based on the assumption of an RS-associated anti-inflammatory effect, these studies should also investigate whether RS supplementation is able to modify the clinical course of PD.

## Methods

### Study design and registration

The interventional study “Effects of resistant Starch on bowel habits, short-chain fatty acids and gut microbiota in Parkinson’s disease” (RESISTA-PD) is a monocentric, prospective, open-label clinical trial investigating the effects of an 8-week prebiotic intervention with the dietary supplement RS (5 g RS twice a day orally) in PD patients (PD + RS) and matched controls (Co + RS). As a third study-arm, PD patients who received solely dietary instructions (PD + DI) were enrolled in this study. Dietary instructions were based on the “Food-Based Dietary Guidelines in Germany” (for further reference see https://www.dge-medienservice.de/food-based-dietary-guidelines-in-germany.html) of the German Nutrition Society. At baseline visit, the specified guidelines to support a health-promoting diet were explained to all subjects in the PD + DI group. These recommendations support a diet rich in whole-grain products and vegetables and a moderate consumption of fat and animal products. Subjects also received a leaflet summarizing these recommendations. This leaflet included a table with practical orientation values for each food group (e.g. cereal products and potatoes; vegetable and salad; fruit; etc.). Primary outcome measures were: change (prior vs. post intervention) in a) bowel habits, b) fecal SCFA concentrations, and c) gut microbiome (analyzed by whole genome-wide sequencing). Secondary outcome parameters were: differences in gut microbiome at baseline (between PD patients and controls), change (prior vs. post intervention) in clinical scales, and change in fecal calprotectin concentrations (prior vs. post intervention).

### Subjects

57 PD patients and 30 control subjects were enrolled. PD patients were assigned to two different interventional groups: PD + RS (n = 32) received 5 g RS twice per day orally over a period of 8 weeks; PD + DI (n = 25) received dietary instructions concerning high fiber intake, but no RS supplementation. Control subjects (Co + RS, n = 30) received 5 g RS twice per day orally over a period of 8 weeks. Main inclusion criteria were an age > 18 years, diagnosis of PD (respectively absence of PD or any other neurodegenerative disorder in the control group), capacity to give written informed consent. Main exclusion criteria were use of antibiotics, steroids, antimycotics or probiotic supplements (during the last 12 weeks), chronic or acute disorders of the gastrointestinal tract (other than constipation), a history of colonoscopy within the past 12 weeks, a history of gastrointestinal surgery (other than appendectomy) within the past three years.

### Clinical assessments

Subjects were assessed at baseline, after 4 weeks and after 8 weeks. Baseline assessment was performed as in-person clinical visit. Assessments after 4 weeks, 8 weeks respectively, were performed as telephone visits. At baseline visit, subjects underwent rating with the Unified Parkinson’s disease rating scale (UPDRS) [33] and the Mini Mental-Status-Test (MMST) [34]. Symptoms related to constipation were assessed at each of the three visits (baseline, 4 weeks, 8 weeks) with the Constipation Scoring System (CSS) [35]. Non-motor symptoms were assessed with the Beck Depression Inventory (BDI) [36] and the Non-Motor Symptoms Questionnaire (NMSQ) [37] at baseline visit and after 8 weeks. In addition to collecting data on adverse events, tolerability and subjective improvement of the intervention was assessed in analogy to the seven-point Clinical Global Impression -Improvement (CGI-I) [38] scale after 4 and 8 weeks. Clinical change was rated (compared to baseline, prior to the intervention with RS respectively) as either very much improved; much improved; minimally improved; no change; minimally worse; much worse or very much worse.

### Collection of fecal samples

At baseline visit, all subjects received sterile containers (P/N S1000-150 and P/N H9550T, Suesse, Gudensberg, Germany) for collection of fecal samples at home. The containers were labeled with the subject-ID and the scheduled time for collection (baseline, i.e. prior to first intake of resistant starch; 4 weeks of resistant starch; 8 weeks of resistant starch). All subjects were instructed how to collect the fecal samples at home and received a leaflet containing relevant information for sample collection. Subjects were instructed to send in two samples (collected on two consecutive days) for each time point. For metagenomics, the first baseline-sample and the first 8-week sample were used, for quantitative analysis of fecal markers. The mean of the two samples was calculated for further statistical analysis. In case of missing 8-week samples, 4-week samples were analyzed (last observation carried forward, LOCF) as described below (see *Statistical analysis of clinical data and fecal SCFA and calprotectin concentrations*). All subjects were reminded by telephone to send in samples after 4 weeks and after 8 weeks. Stool samples were sent to the Institute of Microoecology, Herborn, Germany, and immediately frozen at minus 35° C until analysis.

### Measurement of fecal short-chain fatty acid and calprotectin concentrations

Quantitative analyses of fecal SCFAs and calprotectin were carried out by the Institute of Microoecology, Herborn, Germany. All persons involved in these analyses were blinded to clinical data and the diagnosis of the subjects. Fecal SCFAs were measured by gas chromatography, fecal calprotectin was measured by enzyme-linked immunosorbent assay as previously described [6,18].

### DNA isolation

DNA from fecal samples was isolated using a DNeasy PowerSoil Kit (P/N 47014,QIAGEN, Hilden, Germany) according to the manufacturer’s instructions. To ameliorate the purity, we performed precipitation of the DNA in presence of sodium acetate (pH 5,5) and cold 100 % ethanol at minus 20° C for at least overnight. The DNA was then centrifuged, washed with 80 % ethanol once, and centrifuged another time. The pellet was air-dried and resuspended in TE buffer. DNA concentration was measured using a Nanodrop 2000 spectrophotometer (P/N ND-2000, Thermo Fisher Scientific, Waltham, MA).

### Metagenomic sequencing

DNA libraries were prepared using the MGIEasy DNA Library Prep Kit (P/N 940-200022-00, MGI Technologies, Shenzhen, China) according to manufacturer’s recommendations. In general, 1 µg of input DNA was sheared into fragments using the M220 Focused-ultrasonicator (P/N 500295, Covaris, Woburn, MA). Size selection was carried out using Agencourt AMPure XP beads (P/N A63882, Beckman Coulter, Krefeld, Germany). 50 ng of fragmented DNA were used for end-repairing and A-tailing followed by ligation of barcode containing adaptors to the 3’- and 5’-end. The ligation products were amplified by PCR. A total of 16 different barcoded samples were pooled in equal amount and circularized using a specific oligo sequence, which is complementary to the sequences in the 3’- and 5’-adaptors. DNA nanoballs (DNBs) were generated by rolling circle amplification (RCA), and loaded onto a flowcell using BGIDL-50 DNB loader. Paired-end sequencing was performed according to the BGISEQ-500RS High-throughput Sequencing Set for PE100 on the BGISEQ-500RS instrument (P/N 940-100037-00, MGI Technologies).

### Statistical analysis of clinical data and fecal SCFA and calprotectin concentrations

In case of missing data for 8 weeks and available data for 4 weeks, we applied the last observation carried forward (LOCF) method. LOCF was used for 4 subjects (PD + RS (n = 3) and PD + DI (n = 1)) to replace missing 8-week data concerning fecal markers. Concerning clinical scores, missing 8-week data of 5 subjects (PD + RS (n = 2), PD + DI (n = 1) and Co + RS (n = 2)) was replaced by 4-week data. Normal distribution of data was tested using Shapiro-Wilk’s test. Statistical significance was assumed for p ≤ 0.05. Difference between groups was tested using Mann-Whitney-U-test. Comparisons of the same group at different timepoints was performed with the Wilcoxon’s test for paired samples and sign test for paired samples. Pre-defined outcome measures were not adjusted for multiple testing. Spearman’s correlation coefficient was used to analyze correlations between parameters.

### Sequencing Data Analysis

#### Preprocessing

*FastQC* (version 0.11.8) was used to validate sequence quality and the reports were summarized using *multiQC* (version 1.7) [39]. Adapter contamination was controlled with the *Minion tool* from the *Kraken package* (version 16.098) [40]. None of the samples showed adapter contamination. Trimming and host contamination removal were conducted using *KneadData* (version 0.7.2) (https://huttenhower.sph.harvard.edu/kneaddata/. Accessed 30 Aug 2020) [41].

#### Read-based analysis

Taxonomic composition of the samples was profiled using *mOTUs2* (version 2.5.0) [42] as well as *MetaPhlAn2* (2.9.19) [43]. Both methods are marker-based and were used to profile all taxonomic levels. Functional profiling was conducted using *HUMAnN2* (version 2.8.1) [44]. The *R-package phyloseq* (version 1.28.0) [45] was used to plot the relative abundances in each sample at different taxonomic levels, ranging from kingdom to species. Alpha-diversity was computed using multiple measurements for each sample. The distributions of the alpha-diversity values were compared between patient groups for the same timepoint and between timepoints for the same patient group. Beta-diversity was calculated using the Bray-Curtis distance. Differential abundance analysis was performed by comparing the taxa abundance between groups at the same timepoint and within groups for different timepoints using the R-package *ALDEx2* (version 1.14.1) [46]. *Metacoder R-package [47]* was used to visualize differences in taxa abundance between PD patients and controls. Regression-based balance analysis of the taxa was done using the R-package *selbal* (version 0.1.0) [48]. For analysis with the *selbal* algorithm, we included all samples and all timepoints. Mash distances were computed on the preprocessed reads using *Mash 2*.*1*.*1* [20].

#### Reference-free analysis

Reference-free analysis closely resembled the *BusyBee* workflow [15], which is centered around k-mers. *De novo* assembly was performed using *SPAdes* (version 3.13.1) [49] for all samples with matching baseline and 8-week follow-up dataset. The obtained conigs were filtered by length and sequences shorter than 5000 bases were discarded. Of these filtered sequences longer than 5000 bases, 5-mers and reverse complement 5-mers distributions were computed. Samples were then pooled and a uniform manifold approximation and projection (UMAP) was computed [50]. The embedded datapoints were then reassigned to their respective group-timepoint combination. Contigs for further analysis were taken from the PD+RS group lying within the UMAP coordinates 16.8<X<18,2 and 7.5<Y<11. Remaining contigs were analyzed with *BusyBee*. The reported taxonomic assignment of the filtered contigs was computed with *CAT/BAT* (version 5.0.3) [51].

## Data Availability

Date are available upon request via https://bigd.big.ac.cn/gsa-human/ (project PRJCA004296).

## Supplementary material

### Supplementary figure legends

**Supplementary Figure 1 Alpha-diversity and beta-diversity at baseline**

Supplemental Figure 1 shows measures of alpha-diversity and beta-diversity at baseline between PD patients and controls (**1A** alpha-diversity for *MetaPhlAn2* data, **1B** alpha-diversity for *mOTUs2* data, **1C** beta-diversity for *MetaPhlAn2* data, p 0.001, **1D** beta-diversity for *mOTUs2* data p 0.001).

**Supplementary Figure 2 Intervention-associated changes in clinical scales and fecal markers**

Co+RS: controls plus resistant starch, PD+RS: Parkinson’s disease plus resistant starch, PD+DI: Parkinson’s disease plus dietary instructions **2A** shows the distribution of individual scores in the Constipation Scoring System (CSS), Non-Motor Symptoms Questionnaire (NMSQ) and Beck Depression Inventory (BDI) for the three arms prior to and post intervention. **2B** shows the distribution of fecal calprotectin concentrations in µg per g feces for the three arms prior to (baseline, white) and post intervention (8 weeks, green). For optimized scaling, seven outliers were skipped (n = 3 for PD + RS baseline; n = 2 for PD + RS 8 weeks; n = 1 for PD + DI baseline, n = 1 for PD + DI 8 weeks). **2C** shows the distribution of individual values for the percentage of fecal butyrate for the three arms prior to (baseline, white) and post intervention (8 weeks, green).

**Supplementary Figure 3 Non-metric multidimensional scaling reveals no uniform microbiome shifts associated with the intervention**

Non-metric multidimensional scaling (NMDS) visualizing the microbiome shift due to intervention. As distance measure, the Bray-Curtis measure was applied. Paired data (baseline and 8 weeks) are connected with a segment. PD + RS, Parkinson’s disease patients receiving resistant starch; Co + RS, control subjects receiving RS; PD + DI, Parkinson’s disease patients receiving solely dietary instruction, but no resistant starch.

**Supplementary Figure 4 *BusyBee* Web Results**

Annotated visualization of *BusyBee* Web given the contigs of a differentiated cluster. Each point corresponds to one contig derived from the PD + RS cohort after 8 weeks. If *BusyBee* assigned > 50% of contigs within a cluster to the same genus, the genus annotation is provided.

**Supplementary Figure 5 Pathway Principal Component Analysis**

Principal component analysis results of the center log transformed pathway annotation provided by *HUMAnN2*.

### Supplementary tables

**Supplementary Table 1 Medication of enrolled subjects**

**Supplementary Table 2 Analysis of *ALDEx2* on different taxonomic profiles generated with *mOTUs2***

**Supplementary Table 3 Analysis of *ALDEx2* on metabolic pathways generated with *HUMAnN2***

**Supplementary file: File S1 Supplementary methods**

## Ethical statement

The study was reviewed and approved the ethics committee of the Medical Association of Saarland, Saarbruecken, Germany and registered under the reference number 189/15. The study was registered at the clinical trials registry ClinicalTrials.gov (Identifier: NCT02784145). Written informed consent was obtained from all subjects prior to inclusion in the study.

## Data availability

Date are available upon request via https://bigd.big.ac.cn/gsa-human/ (project PRJCA004296).

## CRediT author statement

**Anouck Becker:** Conceptualization, Formal analysis, Investigation, Writing -Review & Editing, Visualization; **Schmartz Pierre Georges:** Methodology, Software, Validation, Formal analysis, Data Curation, Writing -Review & Editing, Visualization; **Laura Gröger:** Methodology, Investigation, Writing -Review & Editing; **Nadja Grammes:** Methodology, Software, Validation, Formal analysis, Data Curation, Writing -Review & Editing, Visualization; **Valentina Galata:** Methodology, Software, Validation, Formal analysis, Data Curation, Writing -Review & Editing, Visualization; **Hannah Philippeit:** Investigation, Formal analysis, Writing -Review & Editing; **Jacqueline Weiland:** Investigation, Formal analysis, Writing -Review & Editing; **Nicole Ludwig:** Methodology, Investigation, Writing -Review & Editing; **Eckart Meese:** Methodology, Resources, Writing -Review & Editing, Supervision; **Sascha Tierling:** Investigation, Writing -Review & Editing; **Jörn Walter:** Methodology, Resources, Writing -Review & Editing, Supervision; **Andreas Schwiertz:** Investigation, Resources, Writing -Review & Editing; **Jörg Spiegel:** Investigation, Writing -Review & Editing; **Gudrun Wagenpfeil:** Formal analysis, Writing -Review & Editing; **Klaus Faßbender:** Conceptualization, Resources, Writing -Review & Editing; **Andreas Keller:** Conceptualization, Methodology, Software, Validation, Formal analysis, Resources, Data Curation, Writing -Review & Editing, Visualization, Supervision; **Marcus M Unger:** Conceptualization, Investigation, Writing -Original Draft, Supervision, Project administration, Funding acquisition.

## Acknowledgements

This study was funded by the Michael J. Fox Foundation for Parkinson’s Research (Grant ID: 14603) and the German Parkinson Society.

