## Supplemental Figure 1 for "Effects of Resistant Starch on Symptoms, Fecal Markers and Gut Microbiota in Parkinson’s Disease – The RESISTA-PD Trial"

Supplemental Figure 1A

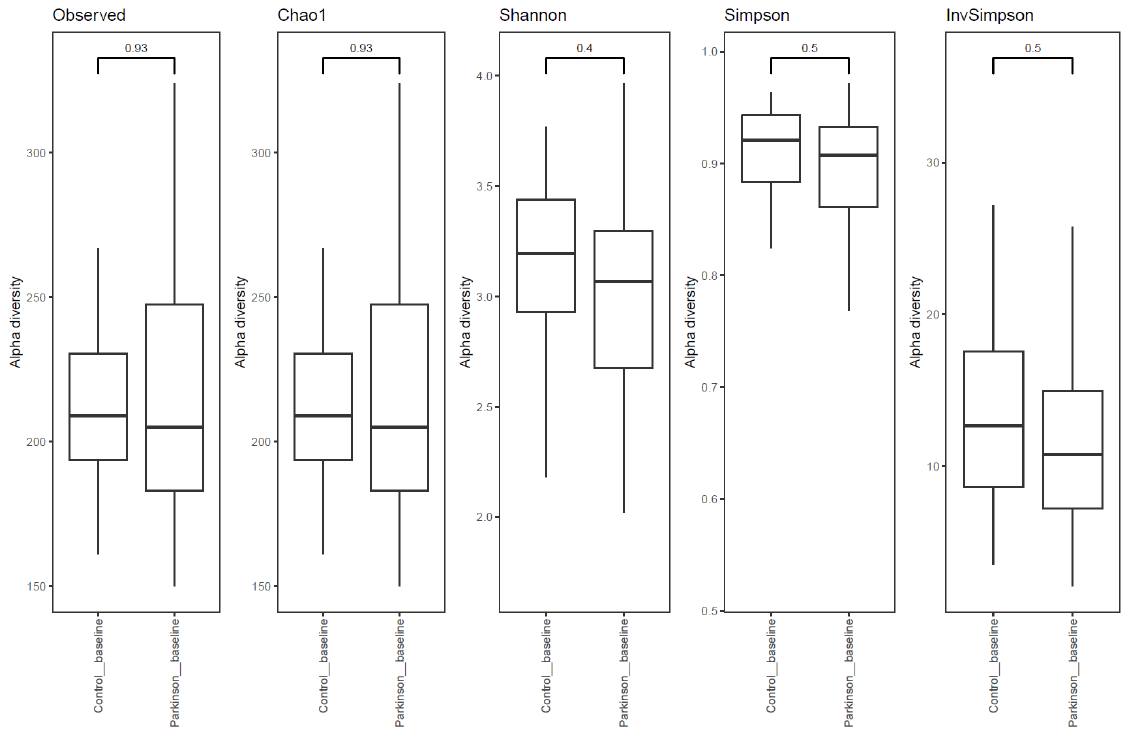

Supplemental Figure 1B

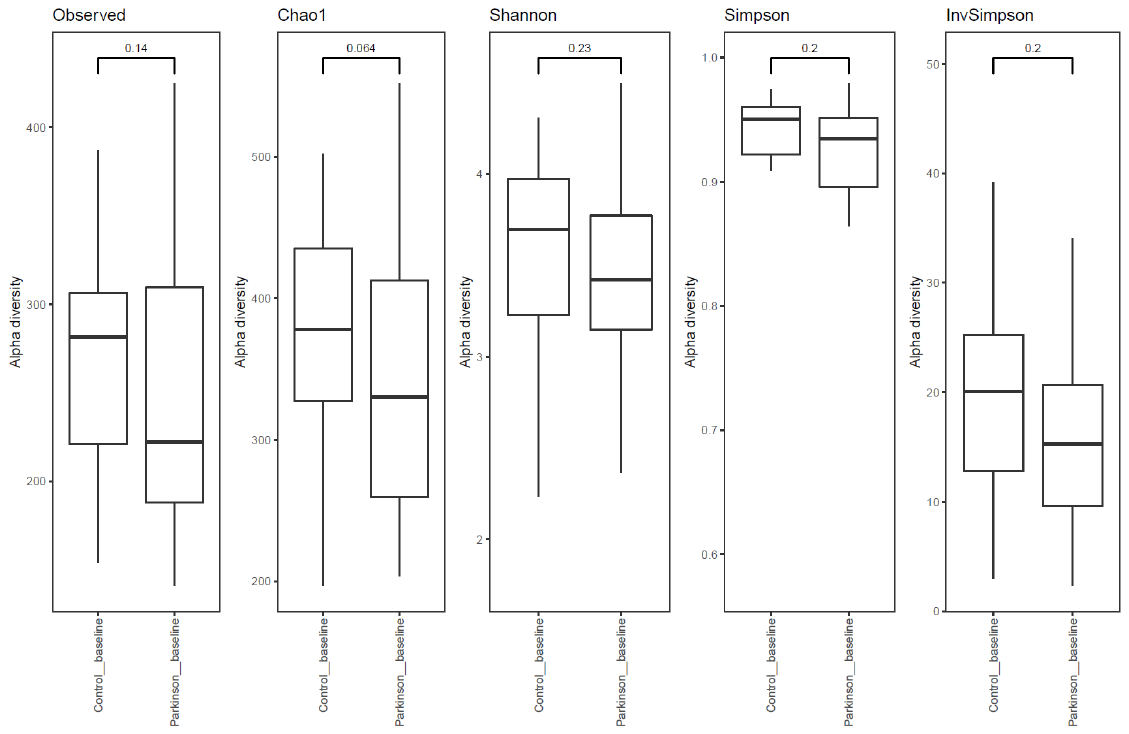

Supplemental Figure 1C

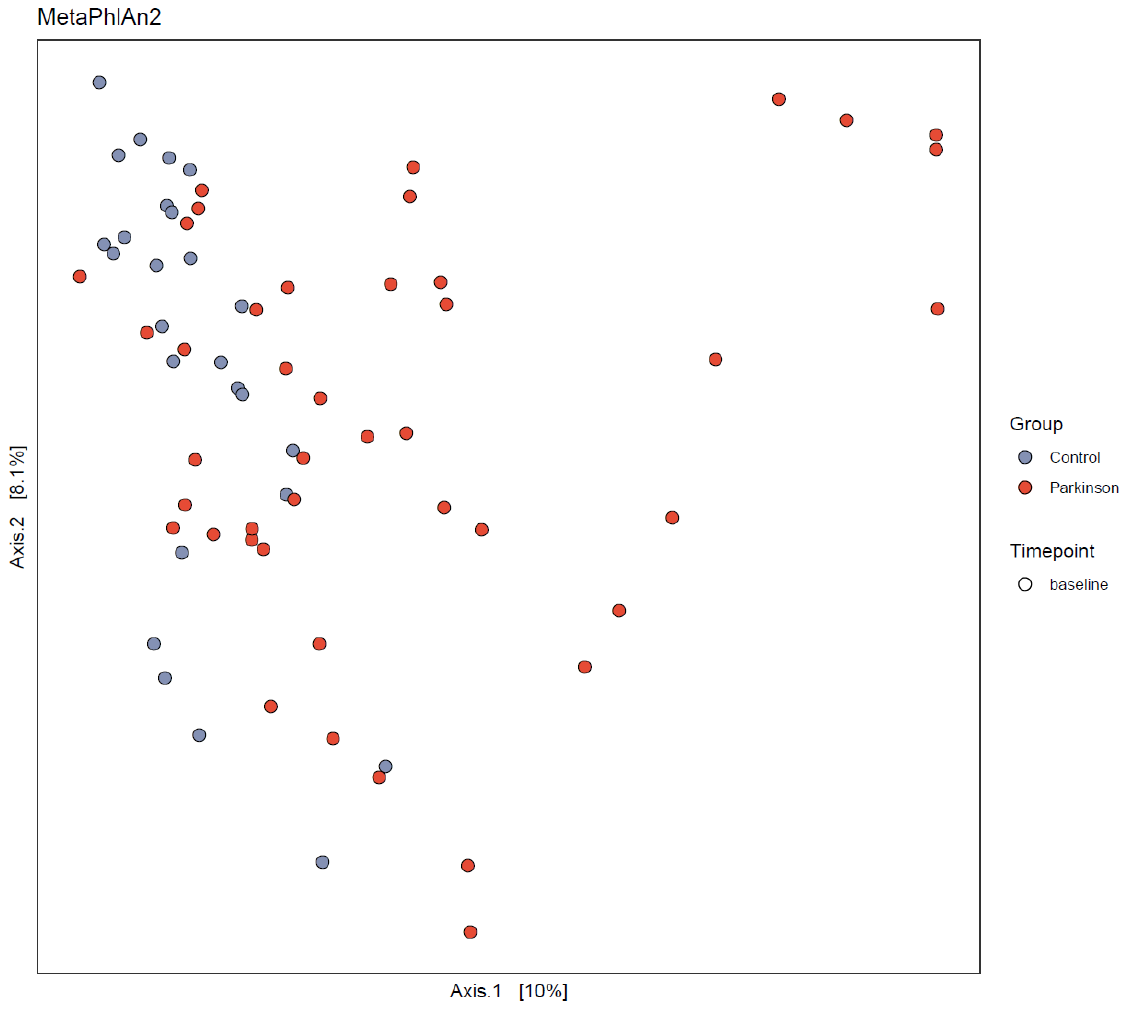

Supplemental Figure 1D

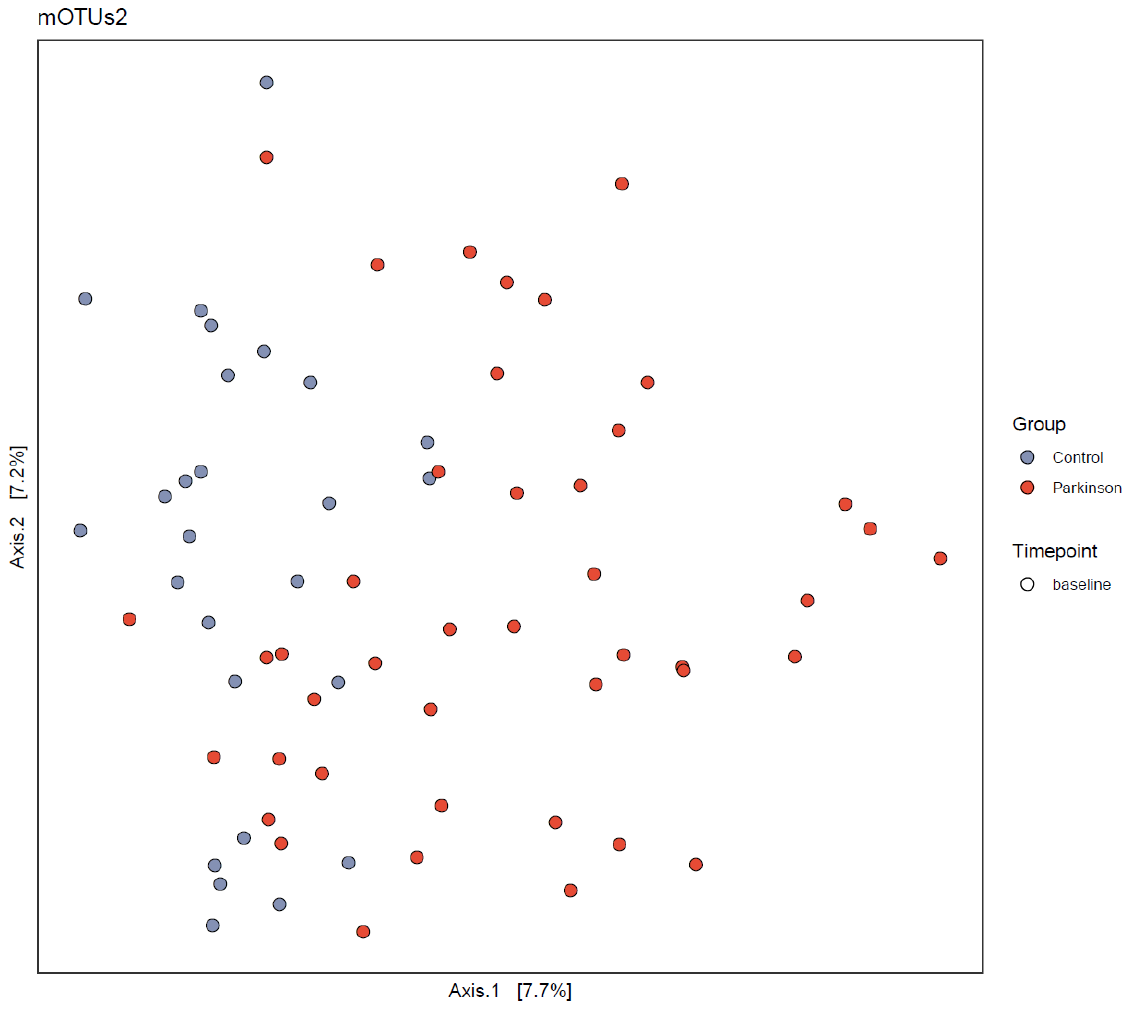

**Supplemental Figure 1** shows measures of alpha- and beta-diversity at baseline between PD patients and controls (**1A** alpha-diversity for MetaPhlAn2 data, **1B** alpha-diversity for mOTUs2 data, **1C** beta diversity for MetaPhlAn2 data, p 0.001, **1D** beta diversity for mOTUs2 data p 0.001).
