## Supplemental Figure 2 for "Effects of Resistant Starch on Symptoms, Fecal Markers and Gut Microbiota in Parkinson’s Disease – The RESISTA-PD Trial"

**Supplementary Figure 2A**

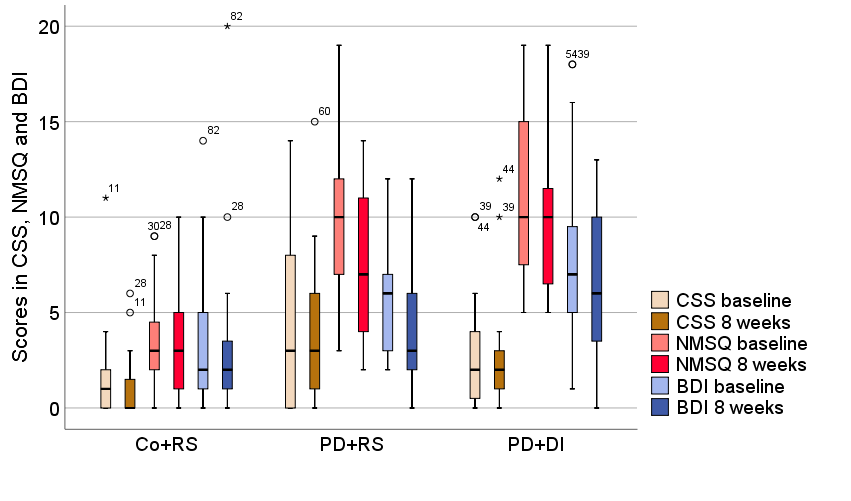

**Supplementary Figure 2B**

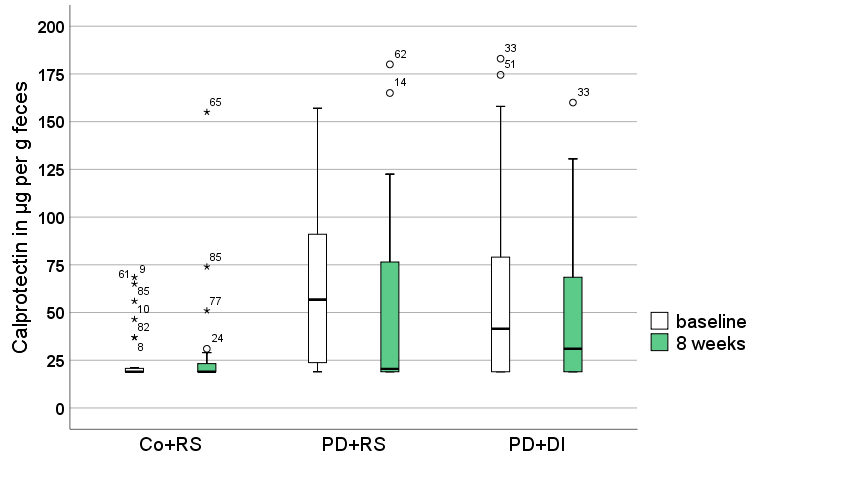

**Supplementary Figure 2C**

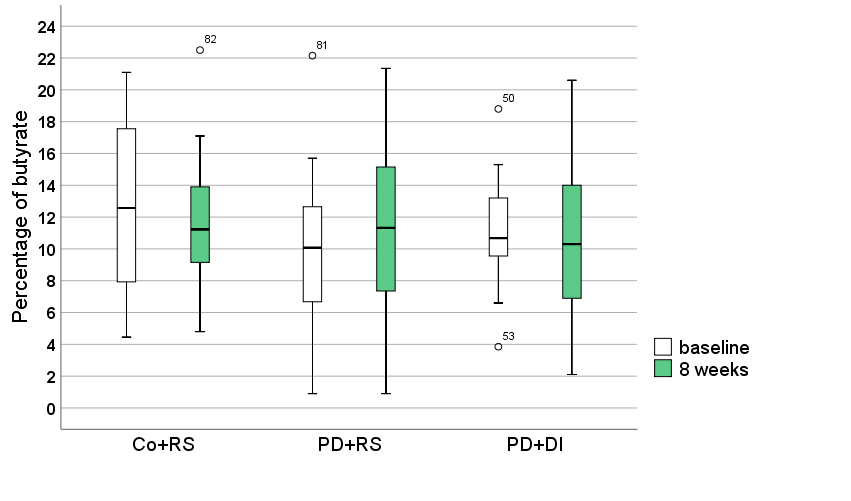

**Supplementary Figure 2 shows intervention-associated changes in clinical scales and fecal markers** (Co+RS: controls plus resistant starch, PD+RS: Parkinson’s disease plus resistant starch, PD+DI: Parkinson’s disease plus dietary instructions)**.** **2A** shows the distribution of individual scores in the Constipation Scoring System (CSS), Non-Motor Symptoms Questionnaire (NMSQ) and Beck Depression Inventory (BDI) for the three arms prior to and post intervention. **2B** shows the distribution of fecal calprotectin concentrations in µg per g feces for the three arms prior to (baseline, white) and post intervention (8 weeks, green). For optimized scaling, seven outliers were skipped (n=3 PD+RS baseline; n=2 PD+RS 8 weeks; n=1 PD+DI baseline, n=1 PD+DI 8 weeks). **2C** shows the distribution of individual values for the percentage of fecal butyrate for the three arms prior to (baseline, white) and post intervention (8 weeks, green).
