## Supplemental Figure 3 for "Effects of Resistant Starch on Symptoms, Fecal Markers and Gut Microbiota in Parkinson’s Disease – The RESISTA-PD Trial"

**Supplementary Figure 3**

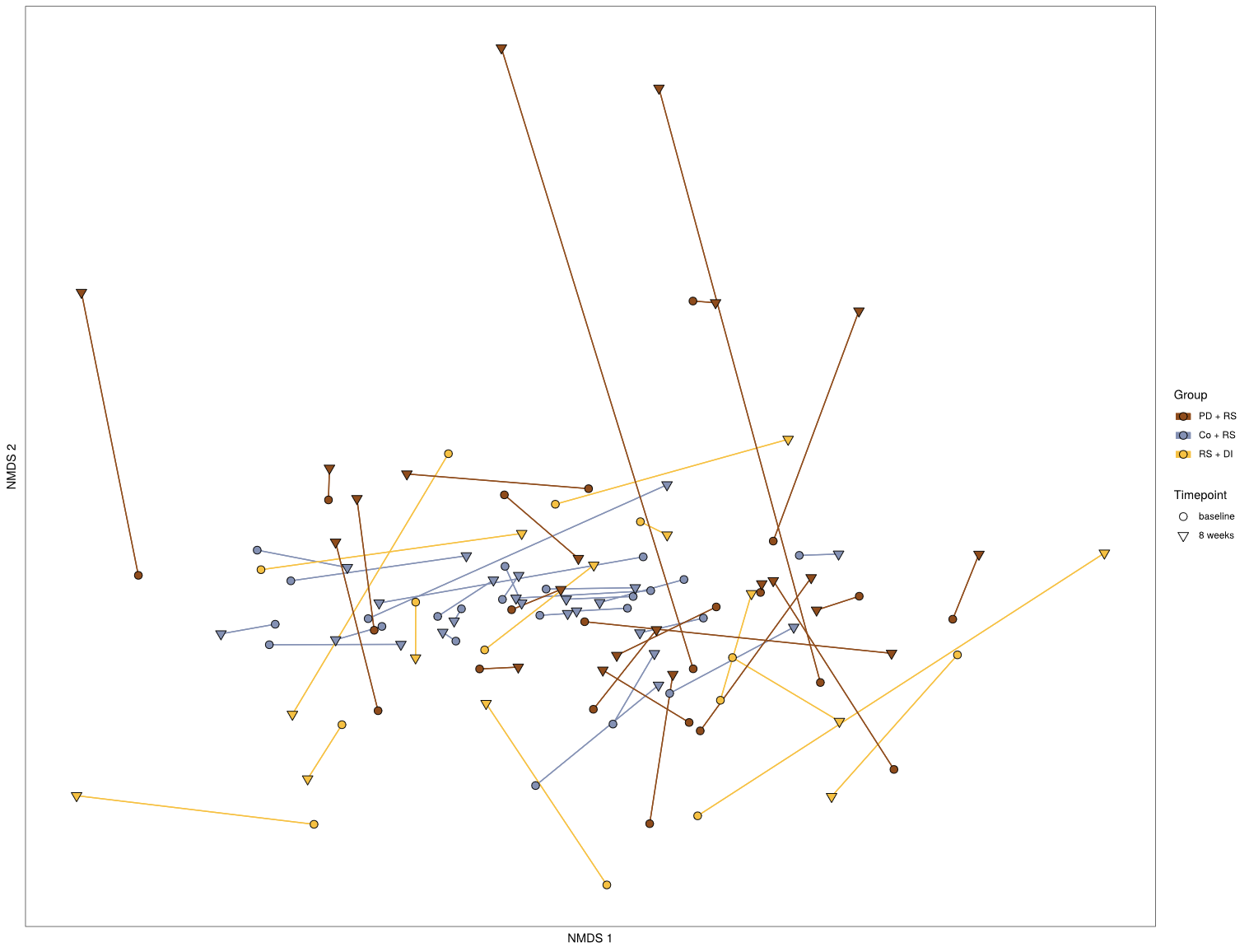

**Supplementary Figure 3**

Non-metric multidimensional scaling (NMDS) visualizing the microbiome shift due to intervention. As distance measure, the Bray-Curtis measure was applied. Paired data (baseline and 8 weeks) are connected with a segment. PD + RS, Parkinson’s disease patients receiving resistant starch; Co + RS, control subjects receiving RS; PD + DI, Parkinson’s disease patients receiving solely dietary instruction, but no resistant starch.
