## Supplementary figures and images for "Effects of Resistant Starch on Symptoms, Fecal Markers and Gut Microbiota in Parkinson’s Disease – The RESISTA-PD Trial"

### Supplemental Figure 4

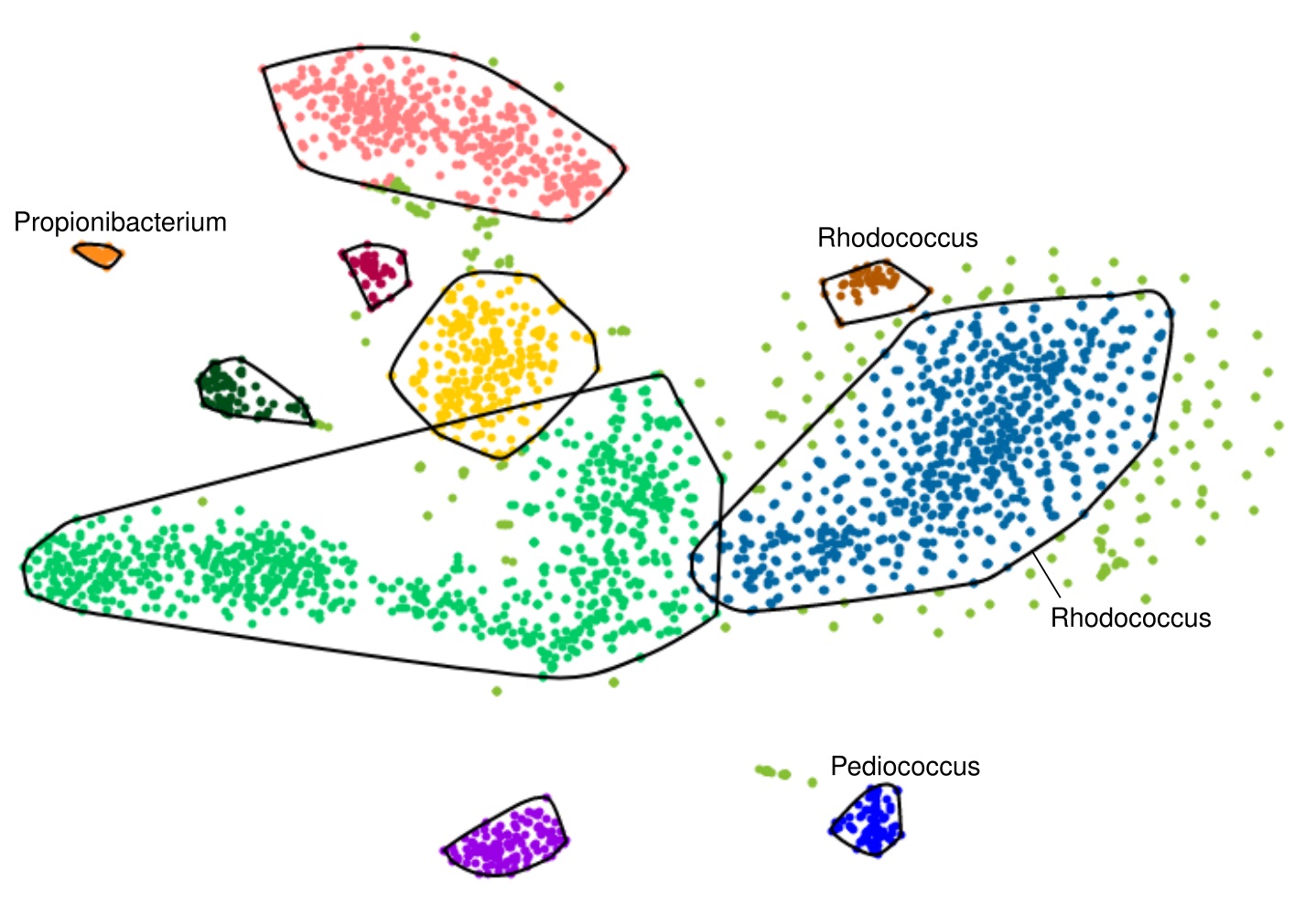

### Supplemental Figure 5

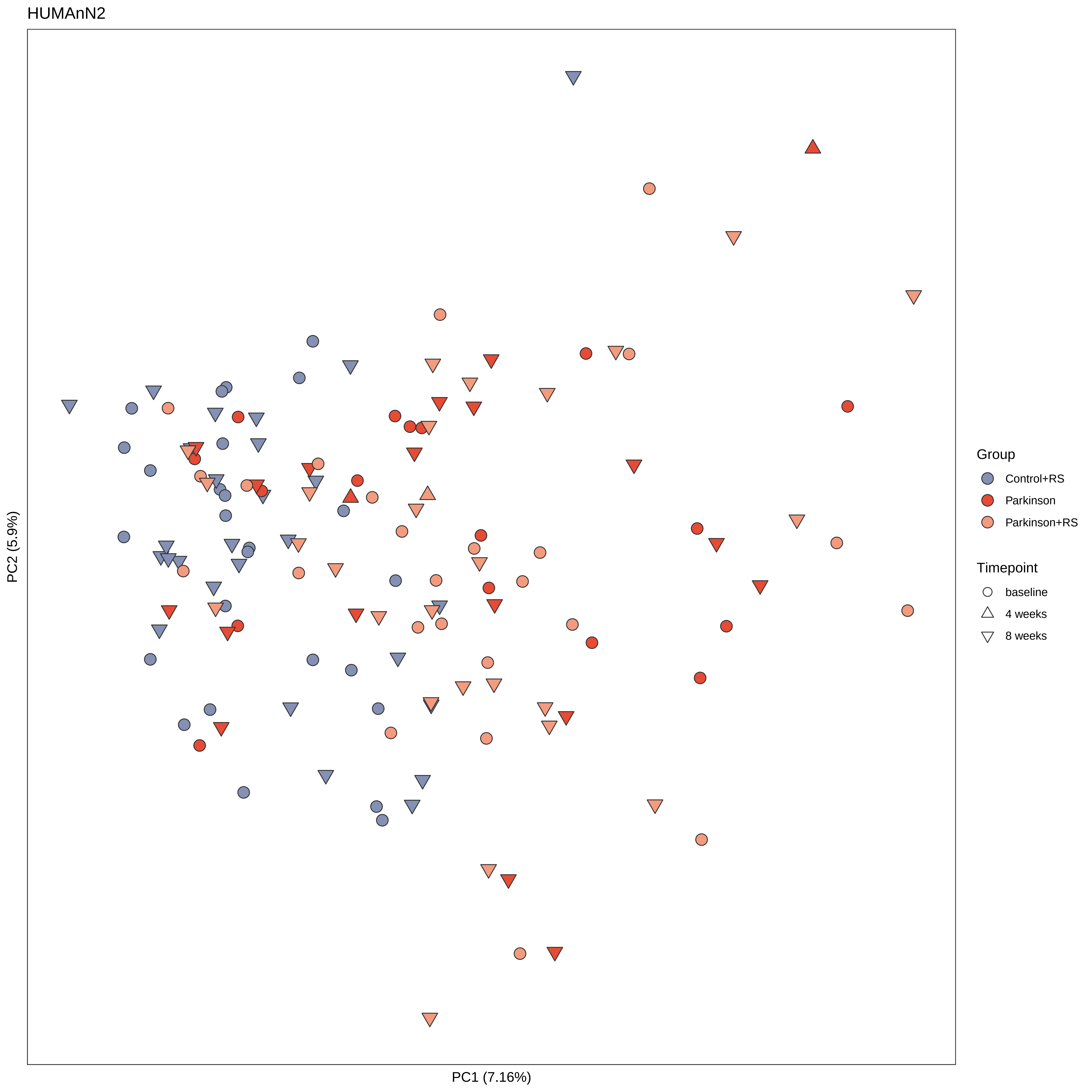
