## Supplemental Table 1 for "Effects of Resistant Starch on Symptoms, Fecal Markers and Gut Microbiota in Parkinson’s Disease – The RESISTA-PD Trial"

### Supplementary Table 1 Medication of enrolled subjects

|  | **Parkinson patients**  **+ resistant starch** | **Control subjects**  **+ resistant starch** | **Parkinson patients**  **+ dietary instructions** |
| --- | --- | --- | --- |
| Number of subjects  enrolled | n = 32 | n = 30 | n = 25 |
| Total daily LED in mg  (median, [range]) | 752 [100 - 1305] | not applicable | 490 [0 - 2010] |
| Therapy with a COMT-inhibitor, type of COMT-inhibitor | 14 of 32, Entacapone: n = 9 Opicapone: n = 3 Tolcapone: n = 2 | not applicable | 8 of 25, Entacapone: n = 4 Opicapone: n = 3 Tolcapone: n = 1 |
| Use of prokinetic drugs | 4 of 32 | 0 of 32 | 2 of 25 |
| Use of NSAID (on demand) | 0 of 32 | 1 of 30 | 2 of 25 |
| Use of proton pump inhibitors | 14 of 32 | 4 of 32 | 12 of 25 |
| Use of laxatives (on demand) | 3 of 32 | 0 of 30 | 0 of 25 |

*Note:* LED, levodopa equivalent dose; COMT, catechol-o-methyltransferase; NSAID, non-steroidal anti-inflammatory drug.
