## Supplemental File S1 for "Effects of Resistant Starch on Symptoms, Fecal Markers and Gut Microbiota in Parkinson’s Disease – The RESISTA-PD Trial"

### Supplementary file S1

**Supplementary methods**

### Read-based analysis

MOTUs2 (version 2.5.0) was run in default setting. MetaPhlAn2 (version 2.9.19) was run to profile the samples in terms of relative abundance (-t rel_ab_w_read_stats) and skip estimation of unknown clades (--unknown_estimation). Afterwards the results for each sample were merged into a single matrix of taxonomy x sample for both tools which was used for further analyses.

Functional profiling was conducted using HUMAnN2 (version 2.8.1) and the results were normalized according to the humann2_renorm_table script, both using default settings. The results for all samples were merged into a single TSV file suitable for further analysis.

The R-package phyloseq (version 1.28.0) was used to plot the relative abundances in each sample at different taxonomy levels, ranging from kingdom to species. Alpha-diversity was computed for each group including the Observed, Chao1, Shannon, Simpson and Inverse Simpson diversity. The distributions of the alpha-diversity values were compared between the same group at different timepoints and different groups at the same timepoint using the Wilcoxon rank test (two-sided, significance level 0.05) (package geom_signif, version 0.6.0). Ordination plots were generated for all samples using multi-dimensional scaling with the Bray-Curtis distance metric.

Additionally, PCA was applied on the count matrices after zero imputation and clr-transformation (by applying the R-packages stats, version 3.5.1 and zCompositions, version 1.3.2.1).

ALDEx2 was run using the Wilcoxon rank test and multiple testing correction with the Benjamini-Hochberg procedure. The significance threshold was set to 0.05 and the effect size threshold to |0.5|.

Selbal was run in linear regression mode using 10-fold cross validation with 100 iterations.

### Assembly-based analysis

SPAdes (version 3.13.1) was run with default settings for metagenomic paired-end data and multiple k-mer sizes (-k 21,33,55). Prokka was run with gene prediction suitable for metagenomes and by ignoring rRNA and tRNA annotation (--metagenome –norrna --notrna). RGI was run with default criteria. Additionally, we searched for the carbohydrate-active enzymes from CAZy database using dbcan2. Finally, functional annotation was done using eggnog-mapper in DIAMOND mode (version 2.0.1) [Huerta-Cepas, Jaime, et al. "Fast genome-wide functional annotation through orthology assignment by eggNOG-mapper." *Molecular biology and evolution* 34.8 (2017): 2115-2122.].

The reads were mapped back to the contigs using bowtie2 (version 2.3.5) with the -no-unal option. Samtools and bedtools were run with default parameters.

CONCOCT was run with default parameters. MaxBin2 was called with -min_contig_length 1500 and MetaBat2 was calle with the --unbinned option and –seed 23. DAS_Tool was called with diamond as search engine. CAT/BAT was run with default options.
